# Randomized vibrotactile fingertip stimulation modulates beta band in Parkinson’s Disease

**DOI:** 10.64898/2026.07.09.26356470

**Authors:** Jesse I. Gilmer, Anthony Lee, Shahin Sharafi, Alexander J. Baumgartner, Thomas K. Uchida, John A. Thompson, Mazen Al Borno

## Abstract

There is growing interest and need for non-invasive stimulation approaches for the treatment of Parkinson’s disease (PD) and other neurological conditions. Pilot studies indicate that vibrotactile stimulation on the fingertips may reduce PD motor symptoms (Pfeifer et al., 2021; Syrkin-Nikolau et al., 2018). PD motor symptoms (e.g., rigidity, bradykinesia) are correlated with exaggerated beta power in the subthalamic nucleus (STN), where neurons are excessively synchronized (Brown 2003; Kühn et al., 2006; Neumann et al., 2016; Yin et al., 2021), but the effect of vibrotactile stimulation on the STN has not been determined. Here, in 12 PD participants in the OFF deep brain stimulation (DBS) and OFF medication state, we investigated how unilateral vibrotactile stimulation applied to the fingertips affects local field potential (LFP) power in STN. We used a within-participants design to expose each participant to a treatment stimulation pattern, termed “Rapidly Varying Sequences” within a Coordinated Reset stimulus framework (“CR RVS”, abbreviated as RVS herein; Pfeifer et al., 2021; Zeitler & Tass, 2015), and a control stimulation pattern, with the order randomized and with intermittent acquisition of STN LFP. RVS yielded a modest but statistically significant 12% (SEM 4.6%) reduction in mean normalized STN beta power and a 48% (SEM 19%) reduction in peak beta power compared to the DBS-off baseline condition and was significantly different when compared to our control stimulus. Furthermore, we identified a biomarker in STN beta power that predicts which participants may benefit from RVS. We observed that participants that exhibited prominent beta peaks had stronger reductions in mean beta power (17% reduction, SEM 6.1%) and peak beta power (55% reduction, SEM 10%). Regressing against the magnitude of the peak in beta provides a moderate prediction of change in mean and peak beta power due to RVS (R2 = 0.58 for mean and 0.52 for peak). We then used our observations to construct a computational model where beta peaks in a simulated STN varied from prominent to diminished. We found that the efficacy of randomized treatments was dependent on the magnitude of beta peaking, mirroring our clinical findings, and showing that RVS may act by reducing intra-neuronal synaptic strengths in STN. Despite robust changes in STN LFP in our study population, we did not observe a significant change in motor symptoms. These results suggest that peripheral vibrotactile stimulation can reduce STN beta power and motivate additional studies to investigate its long-term effects on motor symptoms across a large population of participants.

## Introduction

Parkinson’s disease (PD) is a neurodegenerative disorder that affects 0.5–1% of the population over 60 years of age (Nussbaum & Ellis, 2003). PD is characterized by the loss of dopaminergic neurons in the substantia nigra pars compacta (Bernheimer et al. 1973), which in turn leads to motor dysfunction, including rigidity, bradykinesia, tremor and postural instability (Balestrino & Schapira, 2020; Kühn et al., 2006; Neumann et al., 2016; Yin et al., 2021). PD’s pathophysiology results in abnormal (i.e., excessive) oscillations in the beta band (∼13–30 Hz) of local field potentials (LFP) recorded in basal ganglia cortical loops (Hammond et al., 2007; Ortone et al., 2023). Beta power, when measured in the subthalamic nucleus (STN), is a proxy measure of excessive neuronal synchronization, is correlated with the severity of PD motor impairments (specifically, rigidity and bradykinesia; Little et al., 2012; Feldmann et al. 2021, Kühn et al., 2008), and is inversely correlated with dopamine levels (Radcliffe et al., 2023, Ortone et al., 2023). Deep brain stimulation (DBS) is a frontline advanced therapy for medication refractory PD (Herrington et al., 2016). An increase in DBS stimulation amplitude in the STN reduces beta power (Lofredi et al., 2022; Kühn et al., 2008) and increases upper-extremity movement velocity in patients with bradykinesia (Mishra et al., 2024). However, DBS is an invasive neurosurgical procedure that carries risks, including dyskinesias, dysarthria, and psychological changes (Hariz, 2002).

A potential alternative treatment for PD entails the delivery of mechanical vibratory input (vibrotactile stimulation) via small motors on the fingertips to desynchronize pathological STN networks (Tass 2017; Tass, 2022). Among the earliest observations by Charcot in his investigations into PD was that horseback riding and trips in carriages and trains provided relief from PD symptoms (Goetz, 2011). While whole-body vibration did not prove to be an effective therapy when tested rigorously (Dincher et al., 2019, Kapur et al., 2012), it provided inspiration for stimulation approaches targeting the proprioceptive system that were pioneered by Tass and colleagues (Tass, 2017; Tass, 2022). Meanwhile, DBS strategies were being developed that aim to achieve a long-lasting disruption of abnormal synchrony in the STN, termed “coordinated reset” (CR; Tass, 2003; Adamchic et al., 2014). The delivery of randomized stimuli using vibrotactile sensations was therefore proposed to, non-invasively through sensory pathways, produce the same de-synchronization effects as CR delivered directly to the STN via DBS. Vibrotactile stimulation has yielded clinically promising improvements in PD participants in small clinical pilots (Pfeifer et al., 2021; Syrkin-Nikolau et al., 2018). Previous trials have measured cortical synchrony with EEG (Pfeifer et al., 2021), but, to the best of our knowledge, the effects of vibrotactile stimulation on STN have not been examined. Because beta band power would be an ideal biomarker for measuring the efficacy of vibrotactile stimulus, we sought to validate its utility in a population of persons with PD who had LFP recording enabled DBS implants in STN.

For this study, we developed a vibrotactile stimulation glove at a relatively low cost (less than $50 USD) using off-the-shelf materials (see Fig. 1 A, D right), inspired by the design of Tass and colleagues (Tass, 2022). As in Pfeifer et al., 2021, we chose to stimulate the fingertips because of the large cortical areas that are dedicated to their sensory processing (Penfield, 1937). We delivered vibrotactile stimulation to 12 PD participants while measuring LFPs. Participants were off medication for 12 hours prior to and during the 3-hour experiment and ceased DBS for the duration of the experiment. After DBS was turned off, we included a 30 min wash-out period for the beta responses to stabilize before applying any peripheral vibrotactile stimulation (Temperli et al., 2003; Trager et al., 2016). For participants with bilateral DBS leads (8 out of12 participants), we placed the glove on the hand associated with the clearest beta profile peak, or on the hand contralateral to the implant for unilaterally implanted participants. We applied two stimulus types, one CR stimulation pattern with random alternating stimulation sites (“Rapidly Varying Sequences”; “CR RVS” abbreviated as RVS in this study; See: Fig. 1C; Pfeifer et al., 2021) and one control stimulation pattern with all fingertip sites active simultaneously (“All-on”). We hypothesized that the All-on stimulus would not yield neural desynchronization as it lacks the random arrival of stimuli present in RVS and CR, which is a key feature for inducing desynchronization (Tass, 2003; Adamchic et al., 2014; Khaledi-Nasab et al., 2022). Each stimulus pattern was delivered over the course of 1 hour. LFPs, as assayed through the Medtronic Percept interface via the BrainSense Streaming feature, were recorded periodically before the onset of the stimuli (the “DBS-off” baseline period) and during the application of the stimuli. We chose not to record LFPs continuously for the entire study session to conserve the participants’ DBS battery. A subset of the Movement Disorders Society-Unified Parkinson’s Disease Rating Scale (MDS-UPDRS Part III; Goetz et al. 2008; see Methods: MDS-UPDRS behavioral assessments) examination was performed before each stimulus type and after the conclusion of the experiment to test for changes in motor symptomology. The MDS-UPDRS Part III examinations were performed by an experienced movement disorder specialist (AJB) via video recordings. Side effects and adverse effects were revisited 1 week after the experimental session (Methods: Study procedure). A prior study (Pfeifer et al., 2021) reported a single adverse event related to medication withdrawal, but the participant count was low (*N*=3). In the present study, we were able to directly observe changes to LFP readouts, such as beta power, during the session to assay the effect of the vibrotactile stimulation and thus assess the change in LFPs due to the peripheral stimulation. The order of stimulus application was randomized. We present the experimental schedule for each participant in Fig. 1B. Our primary outcome measure was LFP beta power prior to and after the stimulation intervention. Our secondary outcome measure was the modified MDS-UPDRS Part III score.

**Figure 1.**
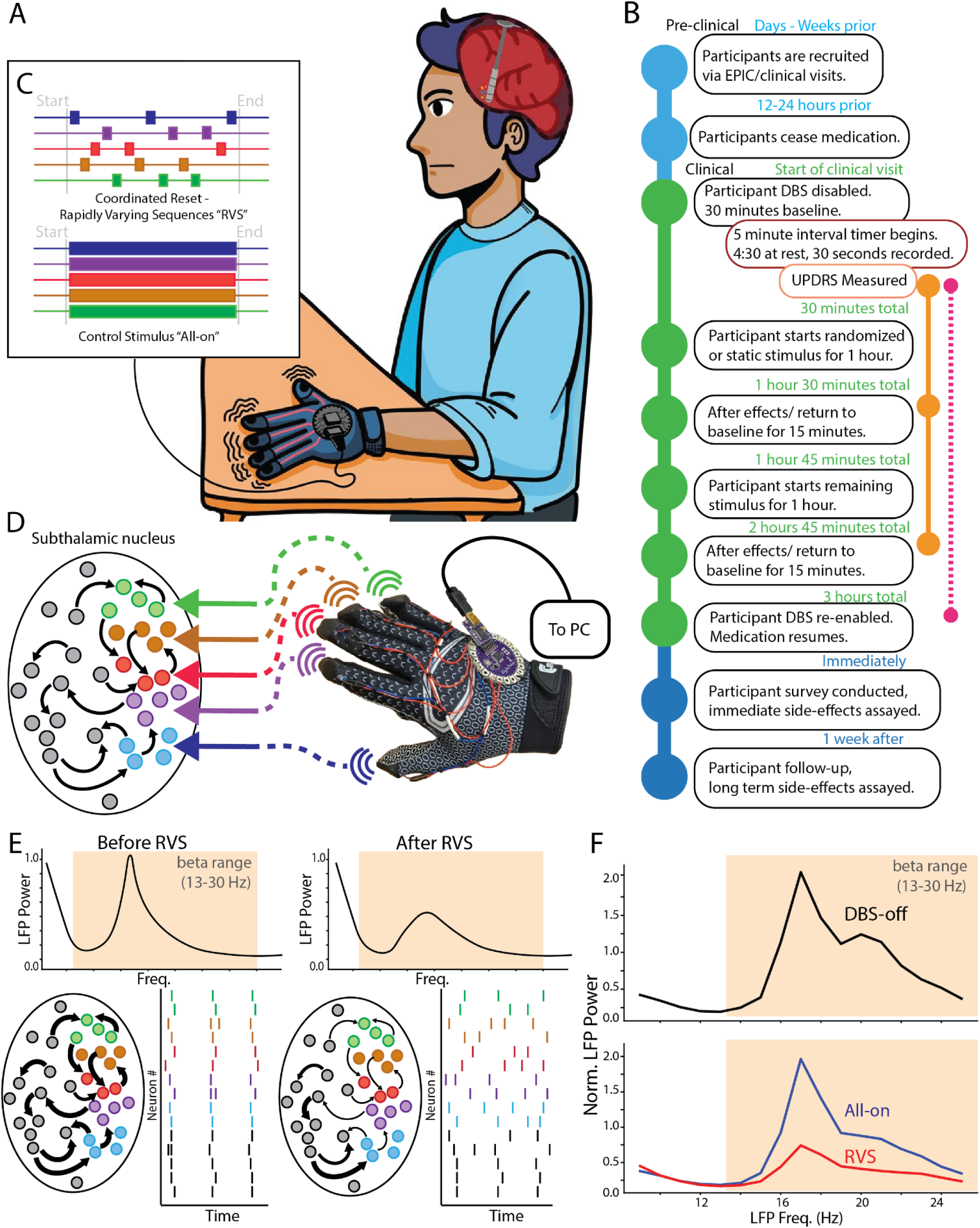
Study design and hypothesis outcomes. **<u>A</u>**: Schematic showing a participant using the vibrotactile stimulation glove. Sensory signals are delivered to the fingertips and the brain responds, influencing the STN through a theorized hyperdirect pathway and brainstem sensorimotor projections. Recordings from the STN are gathered through DBS implants. **<u>B</u>**: Diagram showing the procedure to prepare participants for the study session and undergo vibrotactile stimulation. Cyan dots represent pre-stimulation stages, green dots represent events during the stimulation session, and dark blue dots represent post-stimulation events. The right-sided orange dotted line represents MDS-UPDRS Part III examinations, and pink dots and lines represent recordings from DBS sites. **<u>C</u>**: Two stimulus patterns are delivered: one cycles through random sites on the five fingertips, termed coordinated reset rapidly varying sequences (“CR-RVS”, abbreviated as RVS here; Pfeifer et al., 2021; Zeitler & Tass, 2015); the other (control; “All-on”) targets all five fingertips continuously. **<u>D</u>**: We hypothesize that stimulus from the vibrotactile glove device impacts subpopulations within the STN through sensory pathways. **<u>E</u>**: We further hypothesize that RVS-distributed activity delivered to the STN will reduce overly synchronized activity driven by strong synaptic connectivity within the network. If RVS behaves like coordinated reset (Adamchic et al., 2014; Kromer & Tass, 2024), it will produce an overall stronger reduction in synaptic strength (plots with cells and arrows) leading to less neural synchrony (neural raster plots) which we read out through LFP profiles (top). **<u>F</u>**: An exemplar recording of the beta band in a participant experiencing vibrotactile stimulation with RVS, showing a strong reduction in beta power over baseline (top) and when compared to “All-on” (bottom; RVS is red, All-on is blue).

## Results

### Effects of the stimulation type on STN LFP beta power

Our study included 12 participants with bilateral or unilateral DBS (see Supp. Table 1: Participant Demographics and Implantation Status). 6 participants received the RVS stimulation before the All-on condition, and 5 participants received All-on before RVS. A single participant received only RVS before voluntary cessation of the study due to difficulty tolerating the cessation of DBS. We compared the effects of RVS and All-on on LFP collected from the STN (bilaterally in 8 participants, unilaterally in 4 participants; Fig. 2). For each stimulation type we obtained 12 individual LFP recordings (or 12 “recording sessions” over 1 hour), which had a duration of 30 s and a recording interval of 5 min. We compared the effects of the stimulation types (RVS and All-on) in the final 30 min of each session type to each other and to the DBS-off baseline’s recordings during the final 15 min. Because each participant had a differing range of LFP voltages, we chose to normalize each LFP PSD so that the mean of the analyzed beta band was 0 (see Methods: Local field potential processing). The sampled sessions correspond to the last 30 min of the one-hour stimulations, chosen to give each stimulus type 30 min to act on STN before measuring their effect, which is based on the estimates of CR latency from the computational study by Kromer et al., 2020. We investigated differences in the mean (Fig. 2 A, B, C) and peak (Fig. 2 D, E, F) amplitudes in the power spectral densities (PSDs) across the entirety of beta (13–30 Hz; Fig. 2 A, D), across low-beta (13–20 Hz; Fig. 2 B, E), and across high-beta (21–30 Hz; Fig. 2 C, F), as sub-regions within beta are associated with PD symptomology (e.g. rest tremor with low-beta, Hischmann et al., 2019, rigidity, and bradykinesia, Little et al., 2012; whereas high-beta reflect coupling with cortical structures; Yin et al., 2021) and our participant population had peaking profiles distributed through both low- and high-beta (Fig. 3A).

RVS yielded a statistically significant reduction in the mean and peak beta power compared to DBS-off and All-on (Fig. 2 A, D; <u>mean beta</u>: DBS-off mean = 2.2 percent change [Δ%], standard error of the mean [SEM] = 7.9%, RVS Δ% = -12.1%, SEM = 4.6%, All-on Δ% = -1%, SEM = 4.6%, DBS-off vs. RVS, p = 0.002, All-on vs. RVS, p = 0.003; <u>peak beta</u>: DBS-off Δ% percentage of baseline mean [pom] = 127.1%, SEM = 10.8%, RVS pom = 66.2%, SEM: 5.9%, All-on pom = 112.4%, SEM: 18.8%, DBS-off vs. RVS, p = 0.001, All-on vs. RVS, p = 0.003). The reduction in RVS over DBS-off and All-on was present in peak low-beta, but in mean low-beta RVS was significantly lower than only All-on (Fig. 2 B, E; <u>mean low-beta</u>: RVS Δ% = -9.8%, SEM: 7.4%, All-on Δ% = 4.7%, SEM: 7.4%, All-on vs RVS, p = 0.017; <u>peak low-beta</u>: DBS-off pom = 127.1%, SEM: 19.9%, RVS pom = 66.1%, SEM: 18.8%, All-on pom = 112.4%, SEM: 18.8%, DBS-off vs. RVS, p = 0.001, All-on vs RVS, p = 0.003). RVS was significantly lower than both DBS-off and All-on in mean and peak high-beta (Fig. 2 C, F; <u>mean high-beta</u>: DBS-off Δ% = 4.9%, SEM 10.8%, RVS Δ% = -11.5%, SEM: 4.9%, All-on Δ% = -0.4%, SEM: 4.9%, DBS-off vs RVS, p = 0.0007, All-on vs RVS, p = 0.005; <u>peak high-beta</u>: DBS-off pom = 127.1%, SEM: 19.9%, RVS pom = 66.1%, SEM: 18.8%, All-on pom =112.4%, SEM: 18.8%, DBS-off vs. RVS, p = 0.001, All-on vs RVS, p = 0.003). All-on produced a significant reduction compared to DBS-off only when comparing changes in peak high-beta (Fig. 2F; DBS-off vs. All-on, p = 0.041).

**Figure 2.**
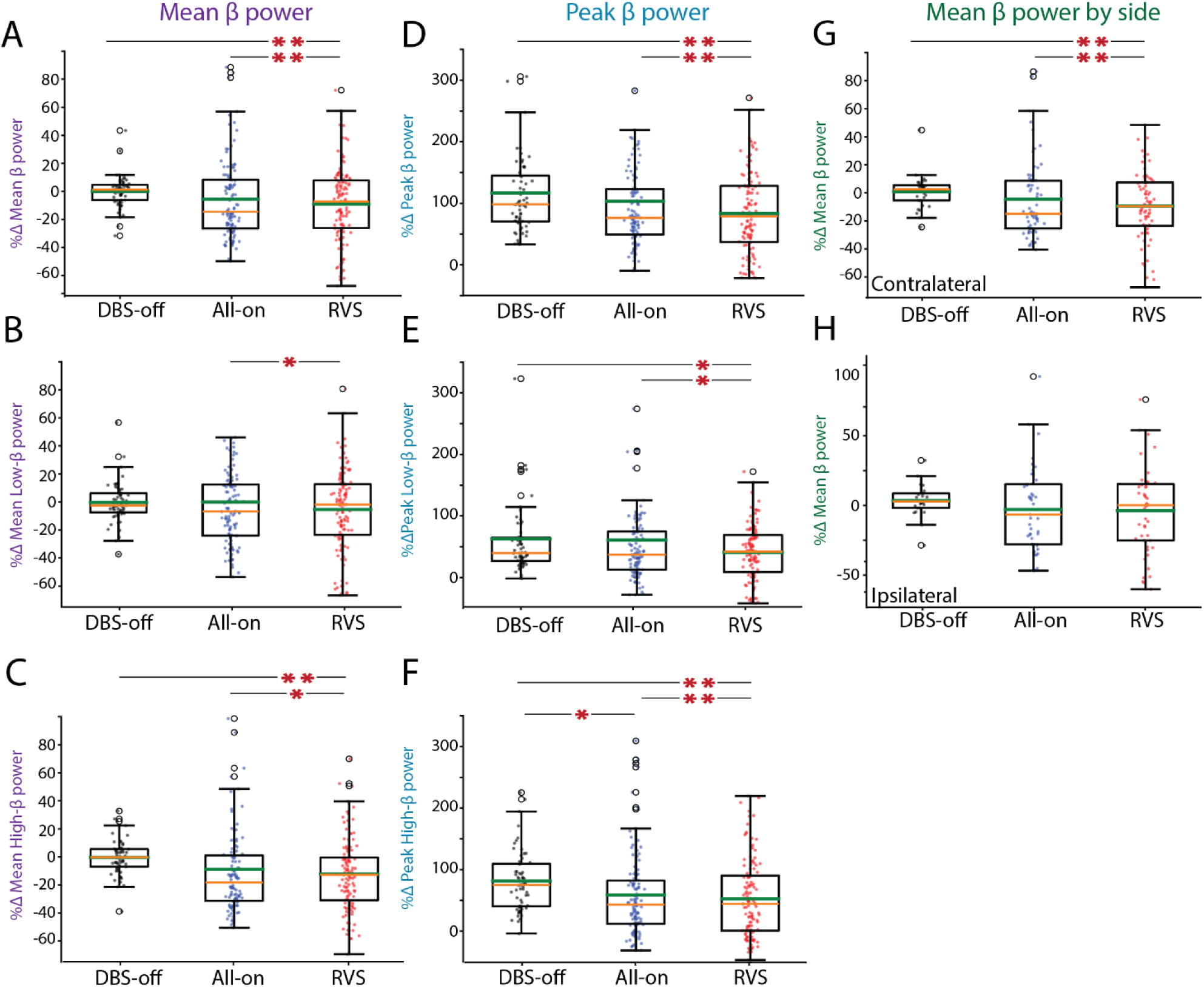
RVS reduces local field potential power mean and peak values. **<u>A</u>:** Comparison of mean “broad” beta power (13–30 Hz) between baseline (DBS-off), the control condition (All-on), and RVS. RVS is significantly different from baseline and All-on (DBS-off mean = 2.2 percent change [Δ%], standard error of the mean [SEM] = 7.9%, RVS Δ% = -12.1%, SEM = 4.6%, All-on Δ% = -1%, SEM = 4.6%, DBS-off vs. RVS, p = 0.002, All-on vs. RVS, p = 0.003). **<u>B</u>**: Same as **A** but restricted to “low” beta (13–20 Hz). RVS is significantly different from All-on but not DBS-off (RVS Δ% = -9.8%, SEM: 7.4%, All-on Δ% = 4.7%, SEM: 7.4%, All-on vs RVS, p = 0.017). **<u>C</u>:** Same as **A** but restricted to “high” beta (20–30 Hz). RVS was significantly different from DBS-off and All-on (DBS-off Δ% = 4.9%, SEM 10.8%, RVS Δ% = -11.5%, SEM: 4.9%, All-on Δ% = -0.4%, SEM: 4.9%, DBS-off vs RVS, p = 0.0007, All-on vs RVS, p = 0.005). **<u>D</u>**: Same as **A** but now comparing the magnitude of peak broad beta power. RVS was significantly different from DBS-off and All-on (DBS-off Δ% percentage of baseline mean [pom] = 127.1%, SEM = 10.8%, RVS pom = 66.2%, SEM: 5.9%, All-on pom = 112.4%, SEM: 18.8%, DBS-off vs. RVS, p = 0.001, All-on vs. RVS, p = 0.003). **<u>E</u>**: Same as **D** but restricted to low-beta. RVS was significantly different from DBS-off and All-on (DBS-off pom = 127.1%, SEM: 19.9%, RVS pom = 66.1%, SEM: 18.8%, All-on pom =112.4%, SEM: 18.8%, DBS-off vs. RVS, p = 0.001, All-on vs RVS, p = 0.003). **<u>F</u>**: Same as **D** but restricted to high-beta. Both All-on and RVS were significantly different from DBS-off, and RVS is significantly different from All-on (DBS-off pom = 127.1%, SEM: 19.9%, RVS pom = 66.1%, SEM: 18.8%, All-on pom =112.4%, SEM: 18.8%, DBS-off vs. RVS, p = 0.001, DBS-off vs. All-on, p = 0.041, All-on vs RVS, p = 0.003). **<u>G</u>**: Same as **A**, measuring the change in mean broad beta power, now only measuring changes in the STN contralateral to the glove hand. RVS is significantly different from DBS-off and All-on (DBS-off mean Δ% = 0%, SEM: 8%, RVS mean Δ%: -15.1%, SEM: 5.6%, All-on mean Δ%: 1.2%, SEM: 5.6%, DBS-off vs. RVS, p = 0.007, All-on vs RVS, p = 0.0004). **<u>H</u>**: Same as **G** but restricted to ipsilateral STN. There are no significant differences. A full summary of statistics of this figure can be found in Supp. Table 3. A, B, C, G, H: Outliers above 100% were not shown for the sake of visual clarity. D, E, F: Outliers above 350% were not shown for the sake of visual clarity. *: p < 0.05, **: p < 0.005. All plots: Orange bars are median, green are mean.

We then compared the effects of the stimulation on the contralateral (Fig. 2G) and ipsilateral (Fig. 2H) hemispheres relative to the hand with the vibrotactile glove. We observed that the reduction in beta power was statistically significant in the contralateral but not ipsilateral hemisphere for RVS when compared to DBS-off and All-on (<u>contralateral:</u> DBS-off mean Δ% = 0%, SEM: 8%, RVS mean Δ%: -15.1%, SEM: 5.6%, All-on mean Δ%: 1.2%, SEM: 5.6%, DBS-off vs. RVS, p = 0.007, All-on vs RVS, p = 0.0004). We controlled for stimulus order by introducing an order parameter into our mixed linear effect model (see Methods: Statistical testing and software) and in no comparison was the order parameter significant (p > 0.05 for all models).

### Baseline beta LFP profiles predict the efficacy of vibrotactile stimulation

On a per-participant basis, both mean and peak LFP beta power was reduced with vibrotactile stimulation (of either type) compared to DBS-off in 9 out of 12 participants for at least one of their DBS leads (i.e., where either RVS or All-on resulted in mean and peak LFP beta power less than 0 after subtracting baseline). While we found that only RVS reliably produced a reduction in beta compared to DBS-off (Fig. 2), the variability of the effects of stimulation on the study population prompted further investigation. Qualitative analysis of participant LFP profiles before and after stimulation suggested that baseline beta profile characteristics may determine the efficacy of vibrotactile stimulation. We classified participants into “peaking” and “non-peaking” beta profile phenotypes and re-examined the effects of vibrotactile stimulation on beta power (see Fig. 3A; see Methods: Beta-peak detection and quantification). When examining the peak frequencies per participant, there was a significantly lower mean beta power RVS stimuli in participants with “peaking” (-P) phenotypes compared to participants with “non-peaking” (-NP) phenotypes (Fig. 3B; RVS-NP Δ%: 9.4%, SEM: 4.3% vs. RVS-P Δ%: -18%, SEM: 6.2%, p = 0.01). This was also true of peak beta power (Fig. 3E; RVS-NP pom: 140%, SEM: 14% vs. RVS-P pom: 53%, SEM: 10%, p = 0.0001), and RVS-P was significantly lower than All-on-P by this measure (Fig. 3E; RVS-P pom: 53%, SEM: 10% vs All-on-P pom: 97%, SEM: 16%, p = 0.039). These results suggest that vibrotactile stimuli was more beneficial for participants with prominent beta PSD profiles and outperforms our control stimulus, All-on, in reducing peak beta. We then analyzed only the 8 participants with a peaking beta profile (5 participants had peaking bilaterally, 1 participant had peaking ipsilateral to the glove, and 2 participants had peaking contralateral to the glove; both contralateral peaking participants had unilateral DBS implants). We compared the last 30 min of the second stimulation (either RVS or All-on) to the last 30 min of the first stimulation. In Fig. 3 C, F, we observe that most participants exhibited a decrease in mean beta profile (i.e. a decrease in the grand mean of either peak or mean of the last 30 min of recording sessions; 6 of 7 STN recordings decreased in peak and 5 of 7 decreased in mean) when RVS was applied on the second stimulation, while some participants exhibited an increase in beta profile when All-on was applied on the second stimulation for measures of mean change in peak LFP (3 of 6 were above 0). RVS 2^nd^ decreased mean beta compared to All-on 1^st^ by a grand average of 14.3% (in terms of percent change relative to DBS-off; SEM: 1.94%; Fig. 3C), and All-on 2^nd^ decreased from RVS 1^st^ by a grand average of 1.4% (SEM: 4.15%) but their distributions did not significantly differ. However, RVS 2^nd^ had a change in mean peak value of -78.1% (Fig. 2F; SEM: 28.8%), and All-on 2^nd^ had a change of +3.1%, (SEM: 11.7%), which was significant (p = 0.044). When examining the last 30 min of recordings (and not their means as in Fig. 3C), RVS as the second stimulation yielded a statistically significant reduction in mean beta power compared to All-on (Fig. 3D, All-on Δ%: 5.8%, SEM: 18.2%, RVS Δ%: -15.6%, SEM: 5.2%, p = 0.0) and in peak power (Fig. 3G; All-on pom: 89.8%, SEM: 25.6%, RVS pom: 53.3%, SEM: 5.2%, p = 0.003).

**Figure 3.**
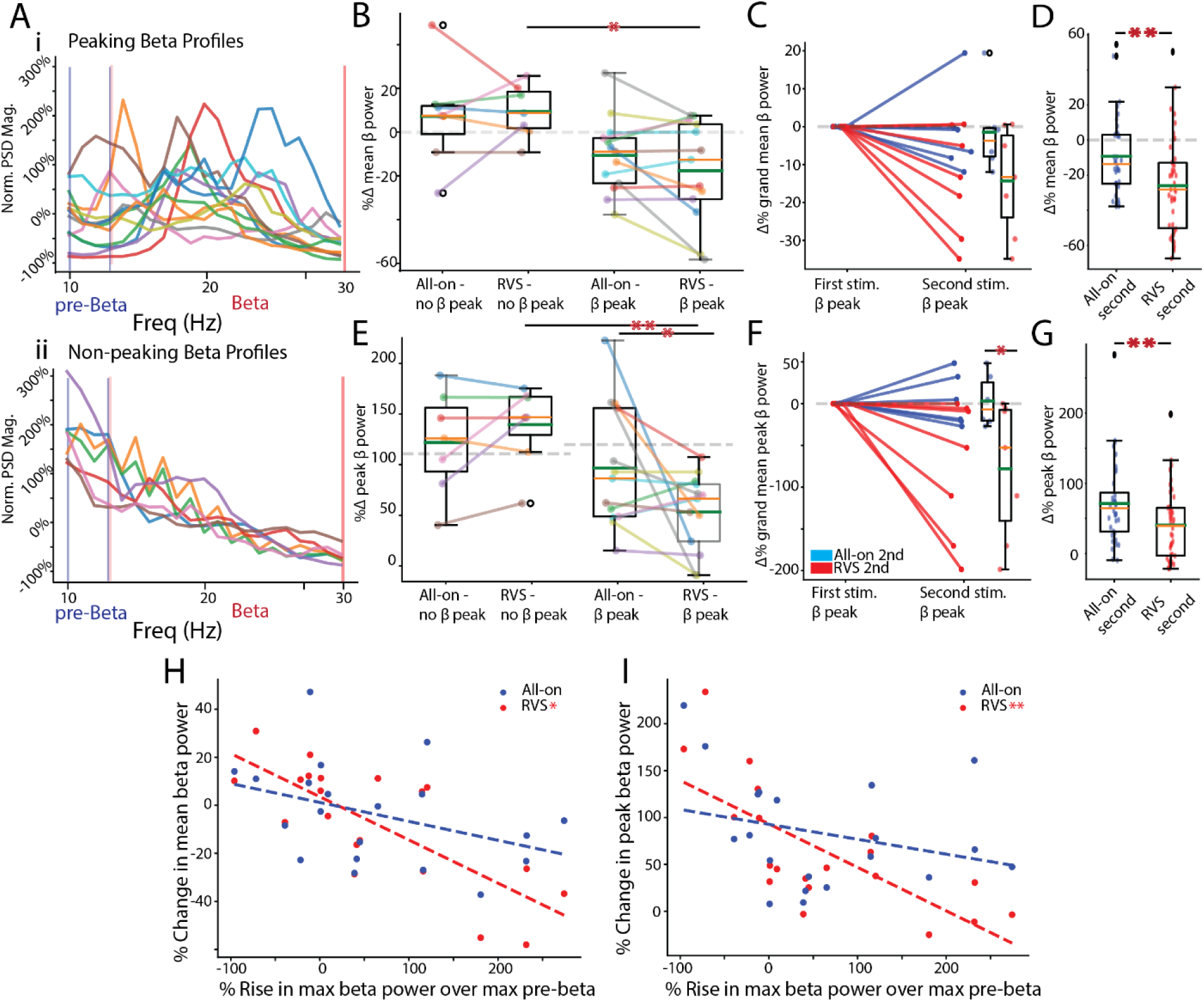
PSD beta profiles predict response to vibrotactile stimuli. **<u>A</u>**: Profiles of 12 participants showing diverse beta-spectrum profiles. “Peaking” profiles show a rise in beta power over the 12 Hz boundary while “non-peaking” profiles decrease from 12 Hz. Red bands show boundaries of alpha and beta frequency ranges. **<u>B</u>**: Decrease in grand mean broadband beta power was significant when comparing RVS between peaked and non-peaked RVS, but not All-on (RVS peaked mean: -17.6%, SEM: 6.18% vs. RVS non-peaked mean: 9.4%, SEM: 4.3%, p = 0.01). Colors of lines are unrelated to colors shown in **A**. **<u>C</u>**: Comparison of effect size on grand means when either stimulus was the second stimulus in sequence. There was no significant difference between RVS and All-on. **<u>D</u>**: There was a significant decrease in the distribution of beta power in the final 6 recordings when RVS was the second stimulus and peaking was present (All-on 2nd mean: 5.8%, SEM: 18.2%, RVS 2nd mean: -15.6%, SEM: 5.2%, p = 0.0). **<u>E</u>**: Same as **B** but comparing peak beta power. Decrease in the participant-specific beta peak value is significantly lower in RVS when there is a peak present (RVS peaked mean: 53.2%, SEM: 10.1% vs. RVS non-peaked mean: 139.6%, SEM: 14.1%, p = 0.0001), and RVS is significantly lower than All-on when a peak is present (RVS peaked mean: 53.2%, SEM: 10.1% vs. All-on peaked mean: 96.7%, SEM: 16.3%, p = 0.039; The percent change in RVS peaked from DBS-off mean = 120% was - 55.6%). Colors of lines are unrelated to colors shown in **A**. **<u>F</u>**: Comparison of effect size when either stimulus was the second stimulus in sequence shows a similar trend as in **C**, but here the grand mean of change in peak RVS is significantly lower than All-on (RVS mean: -78%, SEM: 28.8% vs. All-on mean: 3.1%, SEM: 11.6%, p = 0.044). **<u>G</u>**: As in **D**, the change in peak beta power distributions in the final 30 min of recordings, and when second in sequence, was significant for RVS vs. All-on (All-on mean: 89.8%, SEM: 25.6%, RVS mean: 53.3%, SEM: 5.2%, p = 0.003). **<u>H</u>**: Regression of the rise of max beta over pre-beta (10-12 Hz) value vs. the change in mean beta power in All-on and RVS stimulus conditions. Only RVS had a significant slope (i.e., slope is non-zero) with R^2^ = 0.58 (p = 9e-05). **<u>I</u>**: Same as **H** but regressed against change in peak beta power. Only RVS has a significantly non-zero slope (R^2^ = 0.52., p = 3e-04). A full summary of statistics in this figure can be found in Supp. Table 4. B, C, D, E, F, G: Orange bars are median, green are mean. B, D, E: Grey dashed lines are means for DBS-off. * p < 0.05, ** p < 0.005.

Additionally, we sought to find a relationship between the strength of the beta peak and the magnitude of the reduction in beta power. We defined the strength of the beta peak as the magnitude of the beta band peak relative to the high-alpha band peak (10–12 Hz; frequencies immediately preceding beta were selected to determine if beta band power was increasing over lower frequencies). We performed a linear regression between the strength of the beta peak and the change in beta power following stimulation. We found that change in mean beta power was modestly correlated with the strength of the beta peak when RVS was applied (R^2^ = 0.58, p = 9e-05; Fig. 3H) but change following All-on was not correlated with strength of the beta peak (R^2^ = 0.16, p = 0.08). Change in peak magnitude was also correlated with beta strength when RVS was applied (R^2^ = 0.52, p = 3e-4; Fig. 3I), but again, there was no relationship when All-on was applied (R^2^ = 0.08, p = 0.23). RVS showed a significant correlation with beta strength when examining the contralateral hemisphere for both mean and peak beta, but was only significant in predicting changes in the ipsilateral hemisphere for peak beta (Supp. Fig. 2).

### Effects of vibrotactile stimulation on motor symptoms

We next examined whether vibrotactile stimulation was effective at ameliorating the motor symptoms of PD by calculating changes in the modified MDS-UPDRS (see Methods: MDS-UPDRS behavioral assessments). We examined whether, on average, MDS-UPDRS scores were lower following either stimulation type (Fig. 4A). No significant trend was noted. We then tested for participant-by-participant change in mean MDS-UPDRS score when subtracted from baseline and did not observe a significant change (Fig. 4B). Finally, we separated participants into groups based on their beta peak profiles (see Fig. 3A) and again saw no significant difference (Fig. 4C).

Because participant phenotypes were diverse within the tested population, we reasoned that mean MDS-UPDRS scores may not reflect changes in each participant’s particular motor symptoms. Therefore, we explored differences in individual MDS-UPDRS score distributions (Fig. 4D-E; e.g., change in pronation-supination, finger tapping, etc.; see Methods: MDS-UPDRS Scores) but did not observe a difference in the distribution of individual scores in total (Fig. 4D). Nor did we observe a significant change in the baseline subtracted values (Fig. 4E), nor when separated into peaking and non-peaking phenotypes (Fig. 4G). Further investigation into mean scores sorted by beta profile type (i.e. peaking vs non-peaking) did not exhibit any significant relationship to MDS-UPDRS Part III scoring (Supp. Fig. 3).

**Figure 4.**
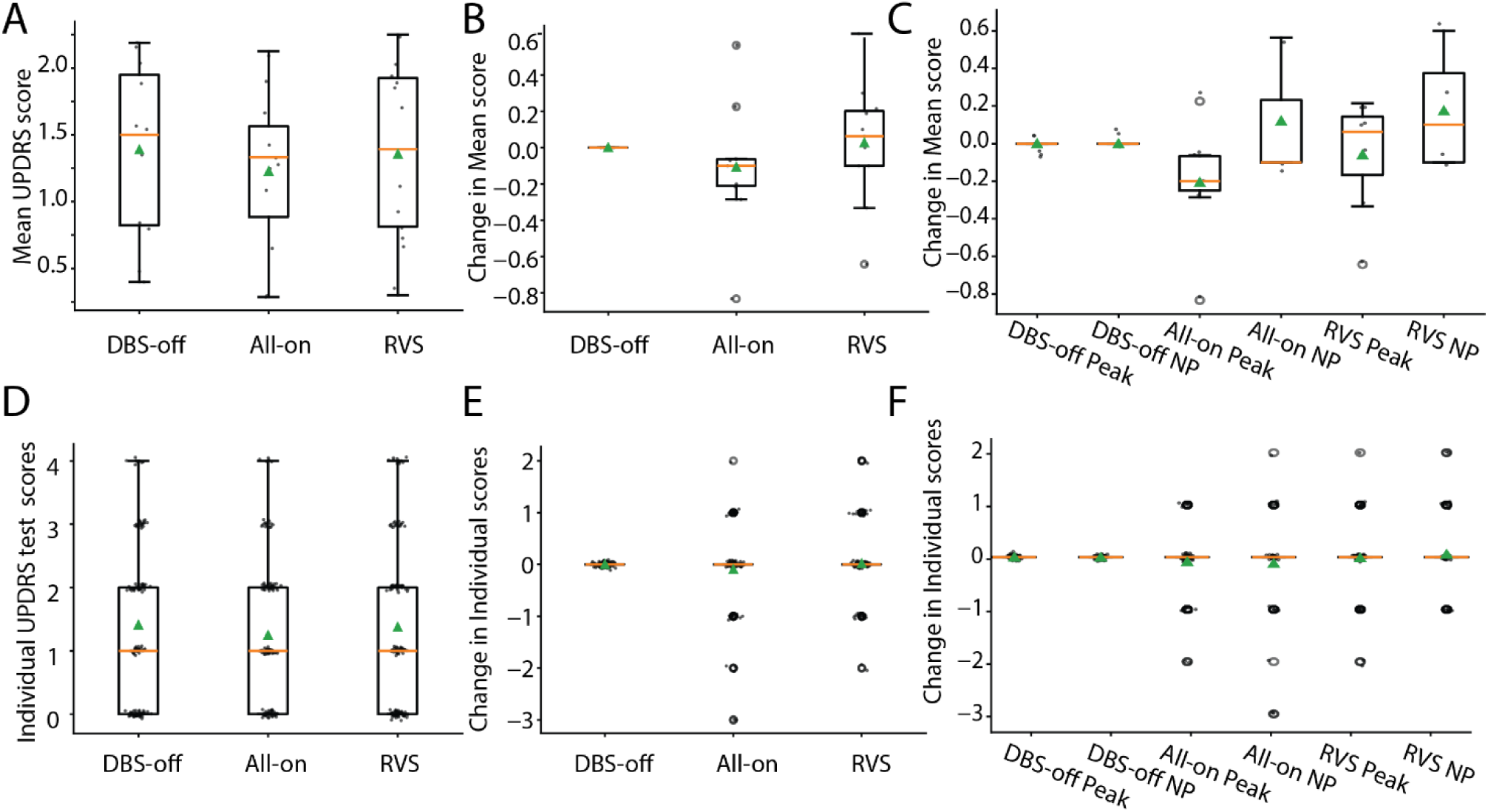
MDS-UPDRS scores were unaffected by vibrotactile stimulation of either type. **<u>A</u>**: Distribution of mean MDS-UPDRS score per participant for DBS-off, All-on, and RVS. There was no significant change following either stimulation type. **<u>B</u>**: A comparison of changes in mean MDS-UPDRS score subtracted from baseline value. There were no significant changes. **<u>C</u>**: Same as **B** but divided into populations with beta peaks and non-peaking power spectrum densities (see: Figure 3). **<u>D</u>**: Distribution of each individual MDS-UPDRS score (finger tapping, pronation-supination, etc.) per participant for DBS-off, All-on, and RVS. There was no significant change following either stimulation type. **<u>E</u>**: Comparison of changes in each score subtracted from their baseline value. There were no significant changes **<u>F</u>**: Same as E but subdivided into peaking and non-peaking power spectrum density profiles. Orange bars are medians, green arrows are means.

### Model of stimuli effects on STN synchrony and plasticity

We next sought to explain the efficacy of RVS at modulating beta band power and the relationship between beta peak magnitude and stimulus efficacy (Fig 3. H, I) using the model of the STN developed by Sharafi et al., 2026. CR and, by proxy, RVS vibrotactile stimulation is thought to act on STN connection strengths by exposing the synaptic connections in the circuit to synchronous inputs distributed among random subpopulations within the structure (Fig. 1D; Kromer & Tass, 2024). Those inputs are thought to then undergo long-term depression (LTD) because of the timing of the inputs interacting with spike-timing-dependent plasticity (STDP; Tass, 2003; Tass, 2017; Ocker and Doiron, 2019), lowering the average strength of STN synapses and reducing synchrony (Chauhan et al., 2024). We therefore modelled differences in synchronous states within the STN (mimicking the variations in beta power profiles seen in the participant population) by altering the variance of firing rates within STN to create more or less prominent beta power peaks (Fig. 5 A, D, G; see Methods: Model of STN with variable beta peak magnitude). We then measured how efficacious CR is within those networks in terms of reduction of beta power, to reflect our clinical results, and on the mean strength of connections within the network in order to demonstrate that beta power and intraneuronal connectivity strength measures are related. We designed our models to produce peaking and non-peaking models (Fig. 5A strong peaking, Fig. 5D moderate peaking, Fig. 5G non-peaking), and then simulated them to reach a steady state, that is, where the mean intraneuronal weights (“<W>”) achieved a stable state (Fig. 5J, first dashed line) in order to establish a baseline <W= measure (here, <W= is normalized to the value at this baseline period). At baseline, the network with the largest beta peak exhibited a strongly synchronous firing pattern (Fig. 5B), while the network without a prominent beta peak had irregular firing patterns (Fig. 5H), reflecting the relationship between beta peak magnitude and synchronization. We then applied a CR pattern alternating between 5 random clusters of neurons for 10 min (Fig. 5J, first to second dashed line) and then measured the change in <W= and beta power magnitude (Fig. 5 A, D, G, light colored lines). We found that the peaking network had a large reduction in <W= and its pattern of activity became more chaotic (Fig. 5C). This also resulted in a large decrease in peak beta power (Fig. 5A, light blue). In the non-peaking model, CR had a small effect on <W= and peak beta power (Fig. 5G, light gray), and the qualitative change in firing was unremarkable (Fig. 5I).

While the results demonstrated by our exemplar models compare well with our experimental results, we sought to establish a quantified relationship between beta peak magnitude and the efficacy of CR, as in Fig. 3H, I. Therefore, we systematically varied the peak magnitude of beta to measure it against change in <W= due to CR by repeating 5 simulations in 10 networks with varying beta peak amplitudes. We found that there was a similar linear fit between the change in <W= and the amplitude of peak beta pre-CR (Fig. 5K change in <W=; R^2^ = 0.59, p = 1e-16) as well as between amplitude of beta peak and reduction in beta peak (Fig. 5L; R^2^ = 0.69, p = 0.0) to those seen in our participants (Fig. 3I). This further demonstrates that CR, and RVS, seem to be most effective at reducing synchrony in networks with large beta peaks, and may be acting by reducing the strength of connections within the STN network.

### Tolerability of stimulus

Following the restoration of DBS at the end of each participant’s visit, we conducted a brief survey of participant experience with the vibrotactile device and stimulation, and solicited their opinion on the suitability of the device for daily use (Supp. Table 2; see Methods: post-experiment survey). 11 of the 12 participants chose to respond to the survey. We asked four questions in total, two regarding any noticeable changes in symptoms per stimulus type (i.e., “Did you notice a change in your motor or other symptoms after the [RVS or All-on] strategy?), one on the comfort of the stimuli, and one on the prospect of daily use.

7 of the 11 respondents (64%) did not notice any difference following the RVS stimulus, 3 (27%) noted improvements, and 1 (9%) noted worsened symptoms. 4 of 11 (36%) respondents did not notice any change after the All-on stimulus, 3 (27%) noted improvements, 3 (27%) noted worsening symptoms, and 1 (9%) noted a mixture of improvements and worsened symptoms. Regarding the comfort of the stimuli, 2 of the 11 (18%) respondents found all stimuli uncomfortable, 1 (9%) found only RVS uncomfortable, and 8 (72%) found both stimuli comfortable. On the prospect of using vibrotactile stimulation on a daily basis, 6 of the 11 (55%) respondents said they could envision using it daily, 2 of the 11 (18%) did not, and 3 (27%) were equivocal on the prospect. Although the responses were mixed, the majority of participants found the stimulus tolerable, and a slight majority were open to daily use of the device (Supp. Table 2).

We report that one participant chose to end the experimental session during the second stimulus delivery due to discomfort from cessation of DBS.

**Figure 5:**
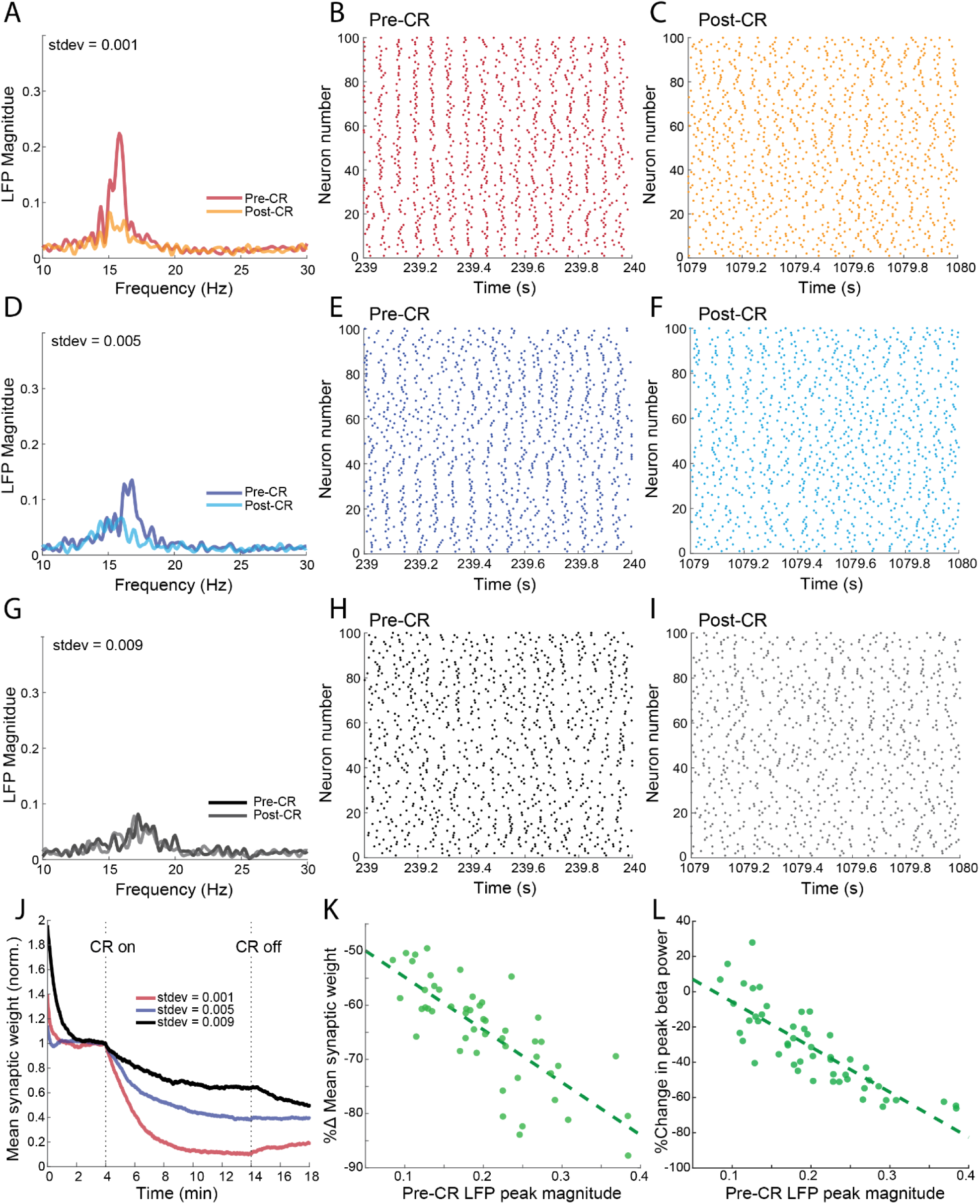
A model of beta peak magnitude’s influence on the efficacy of CR-like therapies. <u>A</u>: Simulated LFP PSDs for a model with a large beta peaking profile. The dark red line is prior to the delivery of CR and the light red line is post-CR. Here, we controlled the variance in firing patterns with a term “stdev” (standard deviation of the term “r”, see Methods: Model of STN with variable beta peak magnitude) which is 0.001 in this model. **<u>B</u>**: A raster of the 100 neurons modelled during the pre-CR epoch showing tightly coupled activity. **<u>C</u>**: Same as **B** but after the cessation of CR, demonstrating desynchronized activity (as hypothesized in Fig. 1E). **<u>D</u>:** Same as **A**, but for a model with moderate beta peaking. The dark blue line is pre-CR and light blue is post-CR. Here, stdev is 0.004. **<u>E</u>**: Same as **B**, but demonstrating less synchrony than the model in **A**. **<u>F</u>**: Same as **C** for the moderate peak model in **D**. **<u>G</u>:** Same as **A**, but for a model with minimal beta peaking. Solid black line is pre-CR and gray is post-CR. Here, stdev is 0.009. **<u>H</u>**: Same as **B**, but with markedly less synchrony than the models above. **I**: Same as **C** for the non-peaking model in **G**. **<u>J</u>:** A plot of the models’ mean weights <W= over the procession of a baseline period (time 0-4 m), delivery of CR stimulation (from time 4-14 m), and a post-CR period (from time 14-18 m). Values are normalized such that the mean weight from time 2-4 m is equal to 1. The red line corresponds to the model in **A**, and the blue line to the model in **D**, and the black line to the model in **G.** Vertical dashed lines denote the onset and cessation of CR stimulation. **<u>K</u>**: Regression of the pre-CR LFP magnitude value vs. the change in mean synaptic weight in the post-CR state compared to pre-CR. We generated 5 simulations per standard deviation of the r term (which varied from 0.001 to 0.010 in steps of 0.001; N = 50). R^2^ = 0.59, p = 6.7e-11. **<u>L</u>**: Same as **K**, but regression is the pre-CR LFP magnitude value vs. change in beta peak magnitude. Note the similarity to Fig. 3I. R^2^ = 0.69, p = 0.

## Discussion

### Effects of vibrotactile stimulus on Parkinsonian brain activity

PD patients may be exposed to iatrogenic effects of dopamine replacement medication (Rascol et al., 2003; Hammond et al., 2007; Aquino & Fox 2014), and DBS, a common second-line treatment, carries its own side effects and risks (Hariz, 2002) that patients and clinicians must carefully weigh. Therefore, non-invasive therapies that can help PD patients manage or lessen the burden of PD symptoms, and their associated interventions, would be beneficial in reducing harm. In this study, we examined the effect of vibrotactile stimulation on PD, namely in the modulation of abnormal increases in power in the beta band (13–30 Hz) of LFP recordings from the STN. Beta band power is a notable biomarker of PD is it reflects an increase in synchronous activity within the STN (Brown, 2003; Kühn et al., 2006; Neumann et al., 2016; Yin et al., 2021; Radcliffe et al., 2023), which are thought to contribute to PD symptoms like bradykinesia and rigidity through the STN’s influence on the globus pallidus and substantia nigra (Blandini, 2001; Benazzouz et al., 2000), and may reflect processing dysfunction in the structure (Brittain et al., 2014). Here, we followed up on the pioneering work of Tass and colleagues in assessing the viability and efficacy of a peripheral vibrotactile stimulation device at reducing PD symptoms. We sought to extend their work by quantifying the effect of the device on the STN by measuring LFPs during the application of the vibrotactile stimulus. Prior studies of vibrotactile stimuli examined changes in MDS-UPDRS ratings of PD symptoms and changes in cortical measures of PD on the brain via EEG (Pfeifer et al., 2021; Syrkin-Nikolau et al., 2018; Tass, 2022), but effects on the STN were not assayed. While MDS-UPDRS Part III scores are a widely used measure of PD motor symptoms, PD patients have heterogenous symptoms that are not always easily measured via behavioral tests. Access to an important biomarker such as beta band PSDs from the STN can help clinicians and researchers assess the effects of this potential therapeutic approach.

### Vibrotactile stimuli reduces beta band mean and peak power in PD

Crucially, this study allows us to test theories that non-invasive peripheral vibrotactile stimulation can produce similar effects to levodopa and DBS on STN beta power (Lofredi et al., 2022; Radcliffe et al., 2023; Ortone et al., 2023). We found that RVS stimulation significantly decreased both mean and peak beta power in our participant population (Figure 2 A, D). While the effects observed in this study across all participants were modest, the stimulus duration was brief in comparison to prior studies (1 hour in the present study compared to 4 hours in Pfeifer et al., 2021) and, as discussed below, reflect heterogeny in participant response to the RVS stimulus.

To the best of our knowledge, a CR-like vibrotactile stimulation pattern has not previously been compared to a control vibrotactile stimulation pattern. This study provides evidence that randomized and multi-focal stimulus patterns may be more effective at reducing STN synchrony than bulk vibrational stimuli (Goetz, 2011; Kromer & Tass, 2024). When examining the control stimulus (All-on), which targets all 5 fingertips simultaneously and continuously, we see that in most comparisons, it did not significantly decrease beta power whereas RVS typically did cause a significant reduction. RVS consistently yielded a significant reduction in the beta band power compared to All-on, suggesting that the effect requires the random, discrete vibrotactile stimulus only present in RVS. Additionally, when participants were grouped into peaking and non-peaking LFP profiles, RVS remained significantly more effective than All-on (Fig. 3 D, E, F, G). While these results are promising, additional evidence that RVS differs from control stimulus patterns requires further research with a larger participant cohort and longer stimulation durations.

### The presence of beta band peaking predicts efficacy of vibrotactile stimuli

Although the change in beta power was significantly different from the DBS-off baseline when investigated across all participants (Fig. 2), the high variance in responses prompted further investigation. Examining PSDs in baseline and after RVS or All-on stimuli revealed that participants had highly heterogenous PSD profiles (Fig. 1F; Fig. 3A). Because high-beta activity is thought to be caused by abnormal synchrony, it follows that a desynchronizing intervention would differentially affect STNs exhibiting high-beta peaks compared to those with flat beta profiles.

Indeed, when we split the participants into those with beta peaks (where beta’s max value rises above that of the pre-beta band) and those without, we found that the effect of RVS was greater in the peaking population. The reduction in mean and peak beta was -17.6% and -55.6% in the peaking population, compared to -12% and -48% in all participants. Furthermore, RVS had significantly lower peak beta values compared to All-on in the peaking population (Fig. 3E). Importantly, again in the peaking population, RVS was significantly more effective than All-on at reducing beta values when applied as the second stimulation in the study (Fig. 3 D, F, G), and treatment order was rejected as a significant contributor to the effects seen in our linear mixed-effects model (p > 0.05 in all tests).

Given that the presence of a beta peak seems to govern the efficacy of vibrotactile stimuli, we then explored whether there was a linear relationship between the magnitude of that peak (defined as the ratio of the peak value to the max value in 10–12 Hz power, that is, immediately before the beta band). When examining the change in mean power following RVS or All-on, we found that peak magnitude was moderately explanatory of the effect of RVS of mean beta (Fig. 3I), and even more so for change in peak beta (Fig. 3H). Intuitively, the more the STN is over-synchronized (i.e., more beta power), the more benefits can be obtained from RVS (i.e., reductions in beta power; Fig. 5). All-on did not have a linear relationship with either mean or peak beta changes following stimulus, providing more evidence that RVS may be more effective than an invariant stimulus not designed to desynchronize neural networks. Finally, the linear relationship with RVS was mainly confined to the STN contralateral to the glove (Supp. Fig. 2), supporting the hypothesis that sensory pathways can modulate STN beta power.

### Beta power peak magnitude may reflect intra-STN connectivity strength and dictate therapeutic efficacy

We concluded our study by modelling a series of neuronal networks with variable LFP beta power peaking profiles (Fig. 5 A, D, G). The variation in patient LFP phenotypes are one of the few metrics available to clinicians that reflect the state of the neural circuits being treated, and the ability to tailor treatment approaches based on LFP may be key to achieving the best possible therapeutic outcome. We sought to link the observed relationship between LFP beta peak magnitude and efficacy of vibrotactile stimulation (Fig. 3 H, I) to a theoretical understanding of CR stimulus’ effect on the neural circuitry in STN as established by the work of Tass and colleagues (Tass, 2003; Tass & Majtanik 2006; Kromer & Tass, 2020). We noted that beta peak magnitude was well correlated with synchronous state in the modelled network (Fig. 5 B vs. C, E vs. F, H and I), which we expected given the well established link between abnormal synchrony, beta band power, and PD pathology (Brown 2003; Kühn et al., 2006; Neumann et al., 2016; Yin et al., 2021; Radcliffe et al., 2023). By having a computational model, we could access measures of intra-neuronal synaptic strength (here termed “weight”), and examine whether CR impacted the mean weight within the STN and whether that change reflected the observed changes in beta peak profiles. We found that CR was most effective at decreasing mean weight when beta power was most prominently peaked in our modelled LFP profiles (Fig. 5 J, K), which was predictive of the outcomes seen clinically with our RVS vibrotactile stimulation intervention (Fig. 3).

### Vibrotactile stimuli and STN modulation may be conveyed through the hyperdirect pathway

Vibrotactile sensory information can modulate STN activity through multiple pathways. The most direct pathway—the hyperdirect pathway—is from end sensory organs of the skin, through the dorsal column–medial lemniscus pathway to somatosensory cortex (Al-Chalabi et al., 2018), which has been shown to project directly to STN through antidromic stimulation studies (Walker et al., 2012; Miocinovic et al., 2018). However, it is unclear whether this is the primary source of neural signaling at work in vibrotactile stimulation or if a more complex, disseminated pathway could be responsible given that STN’s output affects reporting of basal ganglia processing to neocortex. We noted that effects on the STN were significant primarily on the side contralateral to the vibrotactile stimulation glove (Fig. 2G, Supp. Fig. 2). This observation might suggest that the pathway of the CR-like mechanism thought to affect STN is primarily mediated by the hyperdirect sensory pathway rather than by some higher-order or more disseminated mechanism. However, some participants seem to have had both contra- and ipsilateral effects from stimulus exposure (Supp. Fig. 2D), which may lend evidence that there is a “global” benefit of the stimulus.

### Vibrotactile stimulation tolerability

Participant comfort, especially in a potential therapy that would be used daily, is an important consideration. We sought to test a device that could deliver vibrotactile stimulation passively while participants engaged in low-movement activities such as reading or watching television. To investigate whether the device was comfortable, we surveyed our cohort on the tolerability of the device and the prospect of daily use. We also monitored participants for adverse events during stimulation and inquired about adverse events in the week following their study visit. Although the cessation of DBS was characteristically uncomfortable for PD participants, no adverse events or new symptoms unrelated to PD were observed or reported. Overall, a larger-scale study with more participants and longer exposure times is necessary to confirm the safety of vibrotactile stimulation treatments.

Participant reporting suggests that the average experience of vibrotactile stimulation is tolerable, with most participants (Supp. Table 2) not reporting overt discomfort from the stimulus. A majority reported that they would be willing to use the device on a daily basis if it were an effective therapy. However, there were reports of discomfort from the device from some participants which may be improved by adding lighter materials to improve the heat and breathability of the device. Ultimately, determining the appropriate balance between maximizing the effectiveness of the stimulus while minimizing discomfort will be an ongoing aim of vibrotactile stimulation research.

### Study limitations

Although we observed modest, but significant, effects on LFP within the participant population, we did not observe any significant reduction in the modified MDS-UPDRS scores measured after exposure to either the control or RVS stimulus pattern. A prior study into vibrotactile stimulus noted a reduction in MDS-UPDRS scores after an initial 4-hour session (Pfeifer et al., 2021), while our sessions consisted of 1-hour sessions per vibrotactile stimulation type. Given the differences in stimulus duration, we did not expect to replicate the MDS-UPDRS findings shown previously and chose to focus on the impacts on the beta band biomarker as our primary outcome. Although changes in motor symptoms were a secondary outcome of the study, we hypothesize that the accumulation of effects after only 1 hour of exposure was insufficient to produce reductions in MDS-UPDRS scores, even though reductions in beta band synchronization were measured.

A secondary goal of this study was to observe any temporal progression of the vibrotactile stimulus on LFP power, but observation of a timeline of LFP power during the study did not reveal any consistent trends in LFP power (Supp. Fig. 1), and a test of stimulus order in a mixed linear effects model was not significant. Therefore, we did not further investigate how beta power changed over the 1-hour session as the effects of stimulus seemed to appear early and remain consistent across the session recordings for most participants. We ultimately had to balance recording time, validation against an alternative control vibrotactile stimulation pattern, participant comfort, and study feasibility. We found that the total experiment length of 3 hours was well suited to reduce discomfort, as being off both medication and DBS were often physically demanding for participants. This protocol allowed us to recruit participants who were visiting the CU Anschutz Hospital to meet their clinical team on the same day as the study. Future studies may examine the relationship between duration of RVS and the total reduction in beta power and MDS-UPDRS scores following the established efficacy of the RVS in this study.

Another consequence of the 3-hour session duration was a relatively short amount of time for the participants to return to a “PD state” following cessation of DBS. The effects of DBS on PD symptoms are rapid, often alleviating symptoms within minutes or seconds, and the cessation of DBS can similarly exacerbate symptoms within a matter of minutes. The therapeutic effect of DBS on the brain is also thought to diminish within tens of seconds (Kuhn, et al., 2008; Feldmann et al., 2022) to a few minutes (Meissner et al., 2005). Therefore, we reasoned that 30 min would be a sufficient period to allow the STN to return to a PD state while not needlessly extending the duration of the baseline period for participants (Temperli et al., 2003; Trager et al., 2016). We did not observe a stereotyped return to a steady baseline state in the participant population when examining changes in LFP power chronologically (Supp. Fig. 1), and the aftereffects of long-term DBS stimulation are problematic for assessment of putative therapeutic devices. Though patients may return to a PD beta profile upon cessation of DBS, the magnitude of the profile may be attenuated after chronic neurostimulation as measured both 1 minute and 1 hour after cessation of DBS (Trager et al., 2016). Whether this is a new baseline profile for patients following long-term DBS treatment or evidence that the time-course of return to baseline exceeds 1 hour is unknown and may be the subject of further investigation to aid in the design of studies in long-term DBS cohorts.

Conversely, changes in LFP caused by CR are thought to persist for longer durations (Tass et al., 2012), but the time-course of the effect in humans with peripheral CR applications is entirely unknown. Again, we hoped to balance the comfort of our study group with the unknown time-course and chose to take only a brief break between stimulation protocols, typically less than 10 min. Indeed, the overall increasing trend of All-on when administered as a second stimulation may suggest that some medium-term reduction of beta power persisted after the cessation of RVS as a first stimulation (Figure 4; Supp. Fig. 1). While we sought to ameliorate the potential pitfalls of persistent benefit by randomizing the stimulation order, a longer gap between stimulation protocols would be advisable for future research.

Finally, we developed a phenomenological model of the relationship between beta power, efficacy of CR-like therapies, and changes in the intra-STN synaptic strength (Fig. 5). While this model recapitulates the phenotypes we observed within the clinical participant population, (variability in beta profiles seen in Fig. 3A vs. Fig. 5 A, D, G; beta peak to efficacy regressions seen in Fig. 3H vs. Fig. 5K) we emphasize that this model is not meant to exhaustively describe the mechanisms of CR’s effect on neural populations, nor does it attempt to model vibrotactile CR stimulation specifically. We modeled the CR stimulus as a single excitatory pulse, which is not likely to resemble how vibrotactile stimuli alters firing patterns in STN. Additionally, we did not explicitly model any differential pathophysiological process other than changes in firing rates of neurons.

The effects of PD on plasticity both within STN and in the many brain regions impacted by the loss of dopamine remains a key area of research in PD research (Shen et al., 2003). While our results are consistent with the hypothesis that CR primarily functions by decreasing abnormally strong connectivity within STN (Kromer & Tass, 2020) which then drives changes in LFP power (Pfeifer et al., 2021) and firing patterns (Magill et al. 2001), this topic requires further investigation.

### Study Conclusions

Here, we studied the effects of a non-invasive peripheral vibrotactile device on a key biomarker of PD in the STN: beta power. We were able to compare a stimulus pattern, RVS, to a control stimulus, All-on, and demonstrated that RVS is able to significantly lower beta mean and peak power compared to a DBS OFF and medication OFF STN baseline, whereas All-on typically does not (Fig. 2). Further, RVS outperforms the All-on stimulus pattern, showing the efficacy of the random stimulus approach. We next showed that the magnitude of peak beta is predictive of the efficacy of the vibrotactile stimulus, and when comparing the effects of RVS to All-on within a population that has prominent beta peaking, we were again able to show that RVS is significantly more effective than the control stimulus (Fig. 3). These results suggest that RVS-based vibrotactile stimuli may be an effective therapy for managing abnormal STN synchrony. Finally, we developed a neuronal network model of the STN to simulate the variety of beta power phenotypes observed experimentally. This model suggested that a CR-like stimuli, like those that underpin RVS, can reduce abnormally strong synaptic weights and can predict the efficacy of CR based on the magnitude of beta power peaks. This may be an important tool for clinicians to decide which patients may derive a benefit from vibrotactile stimulation therapy and guide further research into peripheral therapeutic device design.

### Future Directions

One striking result of this study was that the profile of the beta band in participant LFP seemed to predict the efficacy of RVS. PD presentation in both symptomology and power spectra is extremely varied. Even within our relatively small population, peak beta magnitude, frequency, and profile breadth were highly heterogeneous. While we applied a stereotyped vibrotactile stimulation to all participants in our cohort, we intentionally designed our device to allow for variation in stimulus frequency, amplitude, timing, pattern, and duration. We hope to further explore the efficacy of RVS parameterization on a per-participant basis to provide the best possible reduction in STN synchronization and reduce the burden of PD symptoms. These objectives may potentially be achieved through the readout of PSDs in patients with DBS implants, through secondary recordings such as electroencephalography (Pfeifer et al., 2021), or through the assessment of MDS-UPDRS scores and motor symptoms. Additionally, we sought to understand the mechanisms underlying the efficacy of RVS in a computational model of the STN. While explanatory models can offer an intuitive understanding of the effects of stimulation, they can also be used to explore optimization of stimulus properties across a variety of PSD profiles to guide parameterization decisions (Sharafi et al., 2026, Khaledi-Nasab et al., 2022), and even to explore dynamic approaches to adjusting stimulation parameters in closed-loop applications. This may be especially useful when modelling the PSD profile specific to each participant, which we explored briefly in this study (Fig. 5).

Furthermore, the selection of the fingertips was based on prior literature that surmised that the dense proprioceptor density of the skin and relatively large representation of the fingertips within sensory regions of the brain make it an ideal location for stimulation targeting (Tass, 2017). While this is intuitively attractive, improving our knowledge of the efferent pathways into STN could improve targeting of stimulation sites. We also suspect that the degree of dispersion or localization of efferent pathways onto the STN makes a significant difference in the amount and degree of synaptic plasticity induced by vibrotactile stimulation. Further computational modelling studies combined with anatomical and functional imaging may provide insight into bi-manual or extra-manual stimulation sites to further improve the modulation of beta band PSDs and PD symptoms.

## Methods

### Participant recruitment

Participants were recruited from the Advanced Therapies for Movement Disorders Center at the University of Colorado Anschutz Medical Campus. Our sample size was chosen based on a power analysis with effect sizes reported in prior work (d=1.001) to provide 80% power based on a paired t-test with 2-sided α level at 0.05 α (Pfeifer et al., 2021). For this study inclusion criteria were the following: between 18 and 85 years of age, have a diagnosis of idiopathic PD, were treated with Medtronic Percept® implantable neurostimulator targeted to the STN, and who could travel to our institution to participate in the study for three hours. Exclusion criteria included patients with severe sensory abnormalities of the fingertips such as vibratory urticaria, patients who were on psychoactive or narcoleptic medications, and patients who were on non-PD medications that affect brain function (i.e., anticonvulsants, attention deficit hyperactivity disorder, depression, or anxiety medication). Participation in our study required that all enrollees refrain from participating in other drug, device, biologic, or intervention trials concurrently or within the preceding 30 days.

In total, we tested 12 participants with advanced idiopathic PD with the Medtronic Percept DBS implanted in the STN. All participants provided written informed consent to the protocol approved by the University of Colorado Institutional Review Board (IRB) #22-1855. Of the twelve participants, one opted out the study session by not completing the second stimulation type. The Medtronic clinician programmer CT900 tablet (loaded with the clinician programmer software A610) and wireless communicator 8880T2 were used to configure the participants’ DBS and conduct the LFP recordings during the study visit.

### Study procedure

The study visit consisted of a single three-hour session at the out-patient clinic of CU Anschutz Clinical and Translational Research Center (CTRC) space. The participants remained seated or laying down (2 lay down, 10 sat) for the duration of the study. Participants were allowed breaks between the baseline and each stimulation recording session. The participants were permitted to use devices (i.e., phones, tablets), listen to music, or read during the treatment (all participants briefly used their phones, one participant watched video on a tablet, and one participant listened to an audiobook). Participants were informed that they could end the study at any time and for any reason, and were periodically monitored and asked about any adverse events. We defined adverse events within the scope of study to be any unintended injury or complication not caused by PD. No adverse events were reported during the study.

DBS impedance was interrogated for each of the three dual-channel configurations for signal health, as well as per implant for dual-DBS participants. Channel configurations with poor impedance were pre-designated for exclusion, but no occurrence of impedance degradation was encountered during the study. Each configuration of channels was inspected after a brief baseline recording with DBS on to identify PSD features, namely the shape and magnitude of beta band (13–30 Hz) peaks. The channel configuration with the highest-powered beta peak was selected for recording. Participants wore one glove on the same hand for both stimulation types. The glove hand was selected based on the best overall channel for examining beta profiles (i.e. by examining LFP on each DBS implant and selecting the lead configuration with the most prominent beta peak).

The sequence in which the two stimulation patterns were applied was determined at random using a simulated coin flip. Following consent, we deactivated the DBS stimulator and started an initial 30 min streaming “session” to establish a baseline. Six 30 s LFP streaming sessions were recorded with a 5 min interval between sessions. We did not record LFPs during the entire stimulation duration to conserve DBS battery. Following the initial six sessions, movements for the MDS-UPDRS were recorded (see Methods: MDS-UPDRS). After establishing baseline (“DBS-off”), one of the two randomized stimulus patterns was applied for one hour with the same LFP recording schedule (twelve 30 s recordings, 5 min interval), followed by the next MDS-UPDRS assessment, a break, and then the remaining stimulation pattern was applied, followed by the remaining MDS-UPDRS assessment (Figure 1B). Participants were then returned to their pre-study DBS configuration and allowed to take mediation if needed.

Participants were observed for 10 min following reinstatement of DBS for instability and other side effects and were contacted 1 week following the study to collect reporting on adverse events and wellbeing.

### Vibrotactile stimulation glove

Gloves were fit with 5 DC motors (part# HD-EMC1003-LW40-R, manufactured by PUI Audio, Inc., ordered from DigiKey.com) with 3D-printed vibration transducers developed in-house. The vibration frequency of the motors was set to approximately 85 Hz (range of 70–100 Hz) by delivering 1 volt via pulse width modulation of 35%. The motors were seated within the fingertips of the gloves, above the fingernail, and vibrations were delivered to the participants’ fingertips through the 3D-printed transducer. The motors were controlled using Arduino LilyPad 328 microcontrollers and powered through a USB cable. The gloves were polyester and cotton gloves made for gardening and produced by a manufacturer named “GloveTacts.” Because of the relatively low cost of the glove components and microcontroller, the glove can be constructed for approximately $50 USD.

The Arduino LilyPad microcontroller was programed to deliver two stimulus patterns: “All-on” in which all five motors were continuously driven, and RVS in which a random sequence of the 5 available motors was selected and then driven for 1 second each before the process was repeated, causing random stimulation across the participants’ fingertips. Methods: Source Code provides the process used to generate the stimulus patterns used in this study.

### MDS-UPDRS behavioral assessments

Participants were asked to perform movements of a modified MDS-UPDRS part III assessment after baseline and each treatment. Movements were recorded with a video camera at 30 frames per second. The tests consisted of Sections 3.4 (Finger Tapping), 3.5 (Hand Movements), 3.6 (Pronation-Supination Movements of Hands), 3.15 (Postural Tremor of the Hands), 3.16 (Kinetic Tremor of the Hands), and 3.17-18 (Rest Tremor Amplitude and Rest Tremor Constancy) of the MDS-UPDRS Part III. Each participant was asked to conduct 10 active movements per hand, at a moderate speed and consistent amplitude, from Sections 3.4, 3.5, 3.6, and 3.16, or hold a posture for 10 seconds in Sections 3.15 and 3.17-18 to the best of their ability. Although not explicitly asked to perform Sections 3.2 (Facial Expressions) and 3.14 (Global Spontaneity of Movement), these metrics were assessed in tandem with other tests. We did not investigate changes in gait. The MDS-UPDRS assessments were rated post hoc by a blinded movement disorder neurologist based on the video recordings (co-author AJB).

### Post-experiment survey

Following the trial and after DBS was restored to a pre-visit state, we asked the participants to respond verbally to a survey with the following questions: “Did you notice a change in your motor or other symptoms after the randomized stimulation [RVS] strategy?”, “Did you notice a change in your motor or other symptoms after the simultaneous stimulation [All-on] strategy?”, “Were the stimulations comfortable?”, and “Could you see this as a day-to-day basis therapy?” The results of the survey can be found in Supp. Table 2.

### LFP processing

LFPs were extracted from the Medtronic tablet in JSON form. Each 30 s recording epoch was assigned a time of recording and participant identifier. For the purposes of examining beta profile features, each band (13–30 Hz for “broad” beta, 13–20 Hz for “low” beta, and 20–30 Hz for “high” beta) was extracted using a bandpass filter (A 7^th^ order Butterworth filter per Hammer et al., 2022). Because each individual participant had an idiosyncratic LFP profile in terms of voltage recorded, we normalized power spectrum densities (PSD) for each individual participant to the last 3 recordings of the baseline period. We normalized such that the mean value across the measured frequency band was equal to 0. Analysis of changes in mean beta power were computed by finding the mean value of the measured frequency range, pooled across all participants, comparing the last 15 min of baseline and the last 30 min of RVS and All-on. Analysis of changes in peak beta power were computed by finding the maximum value within the beta band across all participants, again comparing the last 15 min’s recording sessions from baseline and the last 30 min’s sessions of RVS and All-on. Both mean beta and peak beta were calculated using the percent change formula: (A - B)/B * 100, where A was the condition and B was the DBS-off baseline. In Fig. 3I, and Supp. Fig. 2 B, D change in the peak frequency is defined as change in magnitude of the frequency with the largest value in the first session of the baseline recording (e.g. the 15 Hz band may be the location of peak beta for one participant), which we determined in order to cohere with our definition of peak detection described below. All analyses and transforms were conducted in Python with custom scripts (see: Code Availability).

### Beta peak detection and quantification

Participants presented to the clinic with a variety of beta band PSD profiles. We chose to classify participants as “peaking” or “non-peaking” by comparing the maximum beta value (from 13 to 30 Hz) to the maximum value of the upper alpha band (10–12 Hz). We reasoned that the frequencies immediately preceding beta typically possess a higher PSD magnitude in participants with treated PD (Giannicola et al., 2013), and that PSDs typically follow a “power law” distribution with decreasing power over increasing frequency, so a beta power that surpasses that of the 10–12 Hz range is particularly “peaked”. For each participant, we extracted the PSD from frequencies 10 to 30 Hz, and if the maximum value was at a frequency of 13 Hz or higher, the readings were classified as peaked. To quantify the rise of beta’s peak over the pre-beta band, we calculated the ratio of the beta band’s max value to the maximum value in the 10–12 Hz range.

### Model of STN with variable beta peak magnitude

To model the variable beta profiles observed in our participant population, we utilized a Hindmarsh–Rose (HR) model of the STN that had 100 neurons developed in Sharafi et al., 2026. This model, importantly, is subject to synaptic timing dependent plasticity (STDP; Feldman, 2012) which causes a gradual evolution of intra-STN synaptic strengths to strengthen or weaken depending on the activity profile within the network. While the exact details of the model are available in Sharafi et al. 2026, we highlight here the variables which were tuned to create LFP profiles that resembled the clinical participant population described in this study. Firstly, we adjusted the system of HR equation such that the dominant frequency of the network was within the beta band. Rate was controlled through a parameter called “r” within the HR framework (see: Sharafi et al. 2026 equation 2) . We set r to have a mean value of 0.026, in arbitrary units, and then allowed for a normal distribution around that value by adjusting the standard deviation (“stdev”). A model with a low stdev typically had a very strong beta band peak (Fig. 5A), whereas a model with a large stdev would typically have a very diminished or absent beta peak (Fig. 5G). We then subjected the model to 4 min of simulation to allow the models to achieve a steady-state, both in terms of their intra-neuronal synaptic weights (Fig. 5J) and in terms of firing pattern (Fig. 5 B, E, H). At 4 min we delivered a 4 Hz CR pattern of stimulation. This stimulus was a singular positive current per stimulation site and per cycle, and there were 5 total stimulation sites, reflecting the 5 vibrotactile simulation sites (i.e., the fingertips) used in our clinical experiments.

The progression of stimulation across sites was chosen to be a random permutation of the 5 possible sites for each cycle to achieve an equally distributed but random order. CR progressed for 10 min and then we allowed 4 additional min of simulation before recording the post-CR effect on firing pattern (Fig. 5 C, F, I), mean weight (Fig. 5J), and LFP profiles (Fig 5 ADG; light colored lines). We show example models in Fig. 5A-J, and ran 5 randomized simulations per stdev level, totaling 50 simulations, for Fig. 5 K, L.

Our model used the STDP rule in equation 5 of Sharafi et al., 2026. We used a β value of 2.0, which indicates a system where long term depression has a relatively larger magnitude than long term potentiation. This was chosen to best reflect the beta power peaking observed within our clinical population. The acquisition of LFP within the model was achieved by taking a discrete Fourier transform of the value representing membrane voltage in the HR equations (“x”). Because the sampling rate of the DBS leads used in the clinic was 250 Hz, we achieved a similar level of LFP sampling frequency in our simulated LFP by low passing the signal at 125 Hz.

### Statistical testing and software

We utilized an AB/BA crossover design to randomize our stimulation types. In accordance with guidance on AB/BA crossover study design and to best analyze the effects of stimulation on our outcome measure, beta band power, we used a linear mixed-effects model (LMEM; Lin and In, 2021). We reasoned that LFP power is a noisy measure and therefore used a repeated measures design (30 s recordings obtained every 5 min) to account for variation in recordings, and therefore designed our LMEM model to compare stimulation types to baseline using a standard LMEM model:

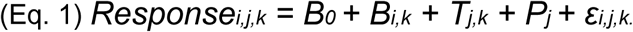

Here *i* is each participant, *j* is the period of the stimulus (e.g. first or second), and *k* is the sequence of stimuli (RVS first vs. All-on first). B_0_ is the intercept of the linear model, B_i,k_ is the random effect intercept for each participant, T_k,j_ is each stimulus (“Treatment”) type with stimulus sequence, P_j_ is the period effect, and ε_i,j,k_ is the residual error. Each participant was treated as a random effect via random slopes (the B_i,j_ term) to allow us to measure within-participant variance (Harrison et al. 2018). The LMEM model was used for comparisons between DBS-off and either RVS or All-on, and a Wald test was used for comparisons of RVS to All-on throughout Fig. 2. A modified LMEM (Effect = B_0_ + B_i,k_ + T_k_) was used for Fig 3. D, G as period term was already controlled for in the data selection. This model was also used for Figure 4 and Supp. Fig. 3.

When comparing mean measures, as in Fig. 3 B, C, E, F, we used independent 2-sample t-tests. When performing regression between rise of beta over alpha, we used a linear regression test and reported the squared Pearson correlation coefficient and the p-value of a Wald Test on the t-distribution (i.e. a test of the hypothesis that the slope is not 0). All statistical tests were performed in Python using the SciPy (Virtanen et al., 2020) and statsmodels (Seabold & Perktold, 2010) packages. Visualizations were generated in matplotlib (Hunter, 2007).

## Data Availability

All data produced are available online at https://github.com/Al-Borno-Lab/Vibrotactile_Clinic_Source_Code_2026

## Code Availability

The code used to generate vibrotactile patterns, analyze anonymized LFP, and generate plots of LFP and MDS-UPDRS summaries, can be found at the following address: https://github.com/Al-Borno-Lab/Vibrotactile_Clinic_Source_Code_2026 For the computational model, please see Sharafi et al., 2026.

## Acknowledgements

JIG designed the study, wrote the manuscript, analyzed LFP and MDS-UPDRS data, performed clinical testing, and administered MDS-UPDRS tests. AL analyzed LFP and MDS-UPDRS data, assisted with clinical testing, and recruited participants for the study. SS designed the computational model. TKU designed the computational model, aided in study design, and supervised the study. AJB assisted with study design, analyzed MDS-UPDRS videos, and provided scoring. JAT assisted with study design, furnished equipment for clinical testing, and supervised the study. MAB designed the study, wrote the manuscript, and supervised the study. This study was funded by a University of Colorado Movements Disorders Pilot Grant to JG, to whom we extend our gratitude. This work was also supported by the Government of Canada’s New Frontiers in Research Fund (NFRF-2023-00258) to TKU and MAB. Sources of funding had no role in study design, data collection and analysis, decision to publish, or preparation of the manuscript.

Thank you to Erin Radcliffe for assistance with analytical practices in DBS and LFP, and for assistance with clinical setup. Thank you to Pamela Gerecht for assistance with participant recruitment. Thank you to Bryce Baker for assistance with clinical setup and MDS-UPDRS recordings. Thank you to the volunteers who participated in our study.

