## Supplementary Materials for "Randomized vibrotactile fingertip stimulation modulates beta band in Parkinson’s Disease"

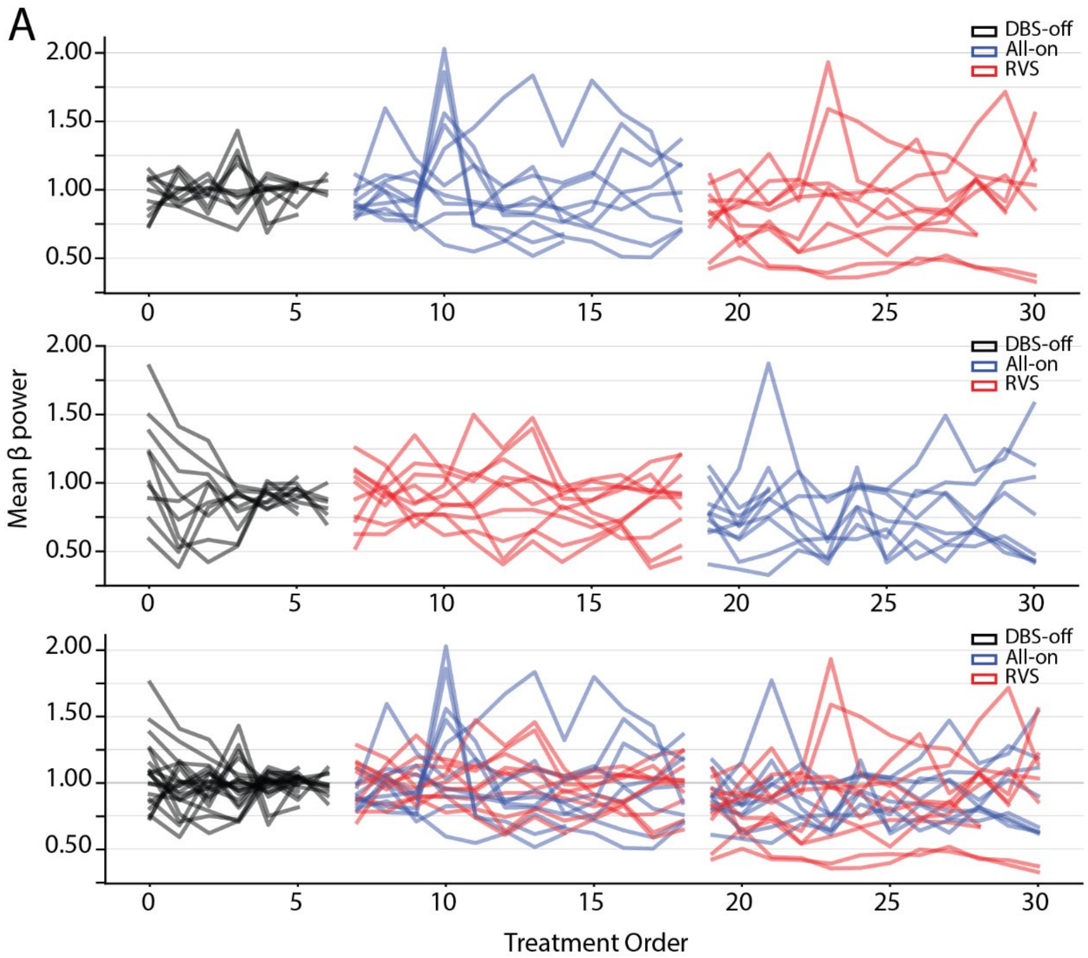

**Supplemental Figure 1. Progression of changes in mean beta power. A:** Time-series showing the mean broadband beta power of each individual participant per recording epoch (i.e. 30 s of recording every 5 min). Red lines are Rapidly Varying Sequences in the Coordinated Reset framework ("CR RVS", abbreviated as RVS) and blue are "All-on". Black lines are the pre-stimulus baselines. All values are normalized so that the last 3 baseline values have a mean of 1.

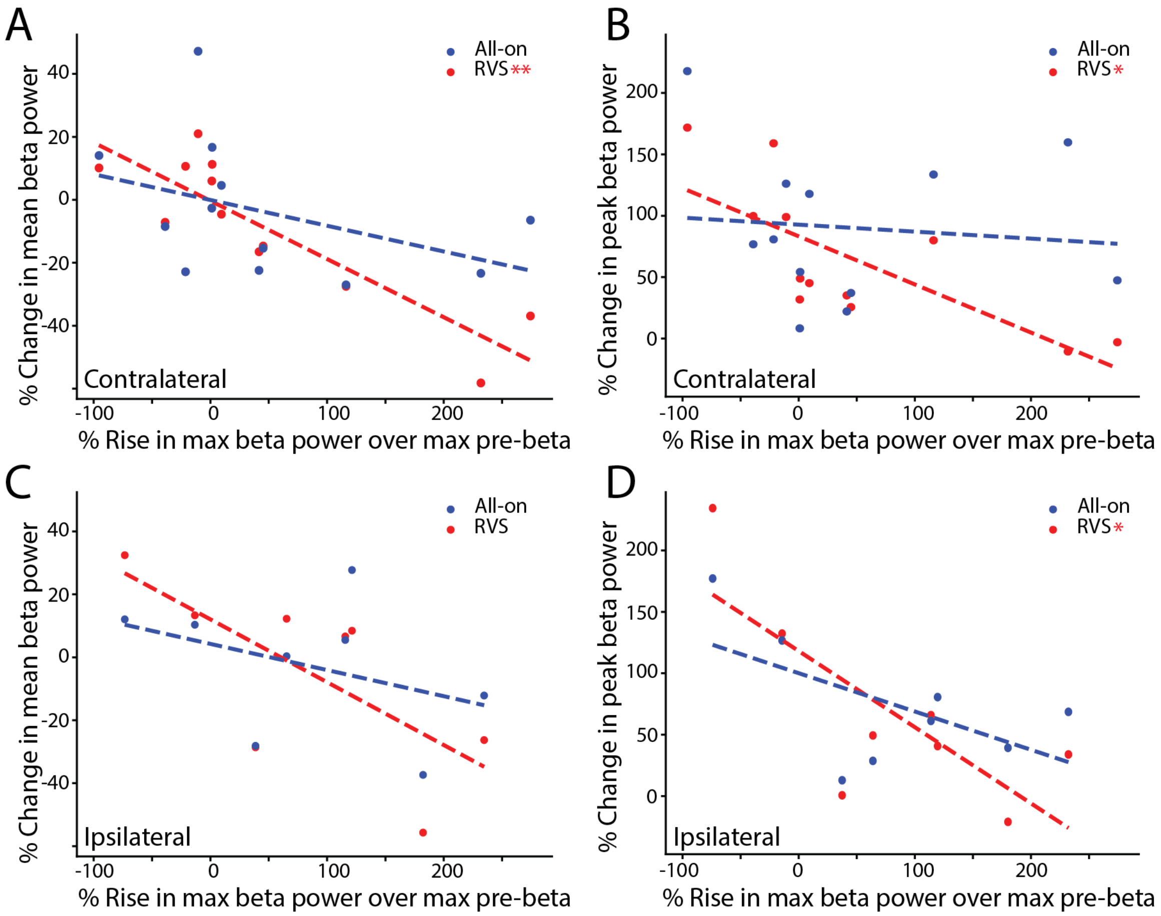

**Supplementary Figure 2. Regression of rise in beta peak against change in mean beta power is localized contralaterally. A:** Regression of the rise of max beta over max pre-beta (10-12 Hz) value vs the % change in mean beta power in All-on and RVS stimulus conditions when examining only the subthalamic nucleus (STN) contralateral to the stimulating glove's hand. RVS has a significant slope (RVS: $R^2 = 0.76$, $p = 0.0002$; All-on: $R^2 = 0.17$, $p = 0.184$)**. B:** Same as **A** but regressing against % change in peak beta power relative to the baseline mean. RVS has a significant relationship to peak beta power (RVS: $R^2 = 0.543$, $p = 0.006$; All-on: $R^2 = 0.01$, $p = 0.76$)**. C:** Same as **A** but for the STN ipsilateral to the glove. There is no significant relationship. **D:** Same as **B**, but for the STN ipsilateral to the glove. RVS has a significant slope (RVS: $R^2 = 0.583$, $p = 0.027$; All-on: $R^2 = 0.33$, $p = 0.14$)**.** * $p < 0.05$, ** $p < 0.005$

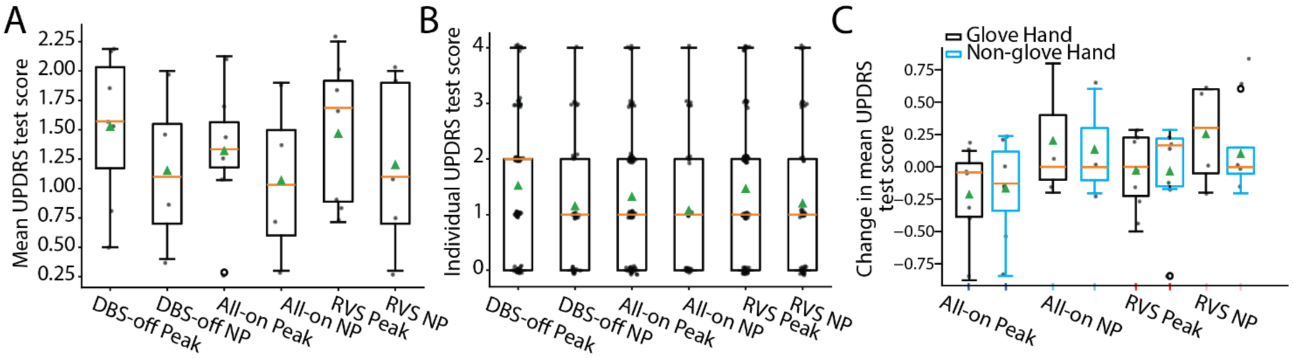

**Supplementary Figure 3. Peaking power spectrum density participants trended towards overall higher MDS-UPDRS scores. A:** Distributions of mean MDS-UPDRS scores per participant following DBS-off as baseline, All-on, and RVS stimulus conditions separated by presence of a beta band peak (see Methods: Beta-peak detection and quantification). Although there is a qualitative trend towards higher mean MDS-UPDRS values for participants with "Peaking" PSD profiles, no significant differences were observed. **B:** Same as **A** but quantified for each individual test score (e.g. finger tapping, pronation-supination, etc.). No significant differences in distributions were present. **C:** Same as **A** but calculated as change in mean MDS-UPDRS score for contralateral and ipsilateral limbs relative to the hand being stimulated and divided into peaking and non-peaking PSD profiles. There was no significant difference between hands per condition.

**Tables:**

| Age (years range) | Biolog-ical sex | Age at PD Onset (Duration, years range) | Age at DBS implant surgery (Duration, years range) | Primary Symptoms | Comorbidities |
|---|---|---|---|---|---|
| 60-64 | M | 45-49 (15-19) | 50-44 (10-14) | Dystonia, dysarthria, gait abnormalities, disordered balance | |
| 65-69 | F | 45-49 (15-19) | 55-59 (10-14) | Bradykinesia, tremor, hyperreflexia, dystonia of foot, gait abnormalities | |
| 55-59 | M | 35-39 (15-19) | 45-49 (10-14) | Ataxia, dystonia | |
| 80-84 | M | 75-79 (5-9) | 75-79 (5-9) | Bradykinesia, dysphonia, dysphagia, gait abnormalities, tremor | |
| 60-64 | F | 50-44 (5-9) | 60-64 (0-4) | Dystonic dyskinesia, bradykinesia, gait abnormalities | |
| 65-69 | M | 55-59 (5-9) | 65-69 (0-4) | Dyskinesia, bradykinesia, dystonia, gait abnormalities | |
| 60-64 | F | 50-44 (10-14) | 60-64 (0-4) | Dyskinesia | |
| 55-59 | M | 30-34 (25-29) | 45-49 (5-9) | Dyskinesia | |
| 60-64 | F | 40-45 (15-19) | 50-44 (5-9) | Gait abnormalities, posture abnormalities, dystonia, tremor | Rheumatoid arthritis |
| 60-64 | F | 50-44 (10-14) | 60-64 (0-4) | Dyskinesia | |
| 65-69 | M | 60-64 (5-9) | 65-69 (0-4) | Tremor, bradykinesia | |
| 75-79 | M | 75-79 (0-4) | 75-79 (0-4) | Gait abnormalities, bradykinesia, tremor | |

Supplemental Table 1: Demographic information of participant population. Here we detail basic demographic information about the 12 participants in this study, showing age, biological sex, age of Parkinson's disease onset with years since diagnosis in parentheses, age of deep brain stimulator implant surgery onset with years since surgery in parentheses, symptoms experienced, and comorbidities.

| **Survey Question:** | **Responses (percent):** |
|---|---|
| Did you notice a change in your motor or other symptoms after the randomized stimulation [RVS] strategy? | Yes, improved: 7 (64%)<br>Yes, worsened: 1 (9%)<br>Yes, mixed: 0 (0%)<br>No: 3 (27%) |
| Did you notice a change in your motor or other symptoms after the simultaneous stimulation [All-on] strategy? | Yes, improved: 3 (27%) Yes, worsened: 3 (27%)<br>Yes, mixed: 1 (9%)<br>No: 4 (36%) |
| Were the stimulations comfortable? | Yes, both: 8 (72%)<br>No, RVS was not:1 (9%)<br>No, All-on was not: 0 (0%)<br>No, neither was: 2 (18%) |
| Could you see using vibrotactile stimulation on a day-to-day basis for therapy? | Yes: 6 (55%)<br>No: 2 (18%)<br>Maybe: 3 (27%) |

Supplemental Table 2: Survey results. Results of a survey conducted with each participant after the experimental session. For methodology see Methods: Post-experiment survey.

Supplemental Table 3: Statistics of Figure 2

| | | | | |
|---|---|---|---|---|
| **2A:** Means, 13-29 Hz (Broad beta), by Treatment Type. | DBS-off Mean: 2.16 DBS-off SEM: 7.87 DBS-off CI: [ -10.79 , 15.11 ] | All-on Mean: -1.05 All-on SEM: 4.57 All-on CI: [ -10.72 , 4.3 ] | RVS Mean: -12.11 RVS SEM: 4.57 RVS CI: [ -21.79 , -6.76 ] | DBS-off vs. All-on: 0.48 DBS-off vs. RVS: **0.0018** All-on vs. RVS: **0.003** |
| **2B:** Means, 13-19 Hz (Low-beta), by Treatment Type. | DBS-off Mean: 0.26 DBS-off SEM: 8.71 DBS-off CI: [ -14.06 , 14.58 ] | All-on Mean: 4.67 All-on SEM: 7.41 All-on CI: [ -7.78 , 16.6 ] | RVS Mean: -9.82 RVS SEM: 7.41 RVS CI: [ -22.27 , 2.12 ] | DBS-off vs. All-on: 0.55 DBS-off vs. RVS: 0.174 **All-on vs. RVS: 0.017** |
| **2C** Means, 20-29 Hz (High-beta), by Treatment Type. | DBS-off Mean: 4.92 DBS-off SEM: 10.82 DBS-off CI: [ -12.88 , 22.71 ] | All-on Mean: -0.37 All-on SEM: 4.85 All-on CI: [ -13.26 , 2.69 ] | RVS Mean: -11.46 RVS SEM: 4.85 RVS CI: [ -24.35 , -8.41 ] | DBS-off vs. All-on: 0.28 **DBS-off vs. RVS: 0.0007** **All-on vs. RVS: 0.005** |
| **2D:** Peak, 13-29 Hz (Broad beta), by Treatment Type. | DBS-off Mean: 127.14 DBS-off SEM: 19.94 DBS-off CI: [ 94.33 , 159.94 ] | All-on Mean: 112.37 All-on SEM: 18.8 All-on CI: [ -45.69 , 16.16 ] | RVS Mean: 66.19 RVS SEM: 18.8 RVS CI: [ -91.87 , -29.02 ] | DBS-off vs. All-on: 0.432 **DBS-off vs. RVS: 0.001** **All-on vs. RVS: 0.003** |
| **2E:** Peak, (Low- beta), by Treatment Type. | DBS-off Mean: 75.23 DBS-off SEM: 18.92 DBS-off CI: [ 44.11 , 106.35 ] | All-on Mean: 69.77 All-on SEM: 18.38 All-on CI: [ -34.68 , 24.77 ] | RVS Mean: 31.04 RVS SEM: 18.38 RVS CI: [ -74.42 , -13.96 ] | DBS-off vs. All-on: 0.77 **DBS-off vs. RVS: 0.016** **All-on vs. RVS: 0.01** |
| **2F:** Peak, (High- beta), by Treatment Type. | DBS-off Mean: 91.47 DBS-off SEM: 19.84 DBS-off CI: [58.84 , 124.1] | All-on Mean: 69.49 All-on SEM: 10.74 All-on CI: [-39.66 , -4.32] | RVS Mean: 43.81 RVS SEM: 10.74 RVS CI: [-64.33 , -29.99] | **DBS-off vs. All-on: 0.041** **DBS-off vs. RVS: 0** **All-on vs. RVS: 0.003** |
| **2G:** Mean, 13-29 Hz (Broad beta), by Treatment Type, Contralateral recordings only. | DBS-off Mean: 0.0 DBS-off SEM: 7.95 DBS-off CI: [ -13.08 , 13.08 ] | All-on Mean: 1.22 All-on SEM: 5.63 All-on CI: [ -8.04 , 10.48 ] | RVS Mean: -14.07 RVS SEM: 5.63 RVS CI: [ -24.33 , -5.81 ] | DBS-off vs. All-on: 0.83 **DBS-off vs. RVS: 0.007** **All-on vs. RVS: 0.0004** |
| **2H:** Mean, 13-29 Hz (Broad beta), by Treatment Type, Ipsilateral recordings only. | DBS-off Mean: 0.0 DBS-off SEM: 11.25 DBS-off CI: [ -18.5 , 18.5 ] | All-on Mean: -9.85 All-on SEM: 7.42 All-on CI: [ -22.05 , 2.35 ] | RVS Mean: -13.08 RVS SEM: 7.42 RVS CI: [ -25.28 , -0.88 ] | DBS-off vs. All-on: 0.18 DBS-off vs. RVS: 0.078 All-on vs. RVS: 0.59 |

Values listed are the results of a Linear Mixed-Effects Model (LMEM; see Methods: Statistical testing and software). Means are the coefficient of regression for the stimulus type added to the intercept (i.e. the coefficient for DBS-off). Statistical tests were LMEM p-values for DBS-off vs. stimulus types or a Wald test for All-on vs. RVS. SEM is the standard error of the mean, CI is the [0.05, 0.95] confidence interval.

Supplemental Table 4: Statistics of Figure 3, S. Fig. 2

| | | | | | |
|---|---|---|---|---|---|
| **3B:**<br>% Change in mean beta power, 13-29 Hz (Broad beta), by Treatment Type and presence of beta Peak | All-on no beta peak mean: 7.25 median: 7.55 st. dev.: 21.66 SEM: 8.187 | All-on beta peak mean: -10.606 median: -8.919 st. dev.: 16.696 SEM: 4.631 | RVS no beta peak mean: 9.413 median: 8.742 st. dev.: 11.38 SEM: 4.301 | RVS beta peak mean: -17.651 median: -12.602 st. dev.: 22.302 SEM: 6.185 | All-on No Beta vs. All-on Beta, p: 0.067<br>RVS No Beta vs. RVS Beta, **p: 0.01**<br>All-on No Beta vs. RVS No Beta, p: 0.83<br>All-on Beta vs. RVS Beta, p: 0.39 |
| **3C:**<br>% Change in the grand mean of beta power, 13-29 Hz (Broad beta), with presence of beta Peak, comparing All-on second or RVS second by subtracting the first treatment. | All-on 2nd: mean: -1.409 median: -3.714 st. dev.: 10.173 SEM: 4.153 | RVS 2nd: mean: -14.291 median: -13.303 st. dev.: 13.087 SEM: 4.946 | | | Change of All-on 2nd vs. Change of RVS 2nd p: 0.1 |
| **3D:**<br>% Change in mean beta power, 13-29 Hz (Broad beta), by Treatment Type, Order, and presence of beta Peak | All-on 2nd Mean: 5.8 SEM: 18.2 CI: [-24.0, 35.7] | RVS 2nd: mean: -14.6 SEM: 5.2 CI: [ -24.1, -7] | | | All-on 2nd, peaked vs. RVS 2nd, peaked **p: 0.0** |
| **3E:**<br>% Change in peak beta power, 13-29 Hz (Broad beta), by Treatment Type and presence of beta Peak | All-on no beta peak: mean: 121.9 median: 126 st. dev.: 47.2 SEM: 17.8 | All-on beta peak: mean: 96.7 median: 86.3 st. dev.: 58.7 SEM: 16.3 | RVS no beta peak mean: 139.6 median146.7 st. dev.: 37.2 SEM: 14.1 | RVS beta peak: mean: 53.2 median: 66.3 st. dev.: 36.5 SEM: 10.1 | All-on No Beta vs. All-on Beta, p: 0.365<br>RVS No Beta vs. RVS Beta, **p: 0.0001**<br>All-on No Beta vs. RVS No Beta, p: 0.49<br>All-on Beta vs. RVS Beta, **p: 0.039** |
| **3F:**<br>% Change in the grand mean of peak beta power, 13-29 Hz (Broad beta), with presence of beta Peak, comparing All-on second or RVS second by subtracting the first treatment. | All-on 2nd mean: 3.118 median: -6.977 st. dev.: 28.54 SEM: 11.651 | RVS 2nd mean: -78.125 median: -52.805 st. dev.: 76.291 SEM: 28.835 | | | Change of All-on 2nd vs. Change of RVS 2nd **p: 0.044** |
| **3G:**<br>% Change in peak beta power, 13-29 Hz (Broad beta), by Treatment Type, Order, and presence of beta Peak | All-on 2nd Mean: 89.8 SEM: 25.6 CI: [47.7, 131.8] | RVS 2nd Mean: 53.3 SEM: 5.2 CI: [33.3, 73.3] | | | All-on 2nd, peaked vs. RVS 2nd, peaked **p: 0.003** |
| **3H:**<br>Regression of change in % change of mean beta power by rise in peak beta over pre-beta. | All-on Regression: p-value: 0.086 R: -0.393 $R^2$: 0.159 | RVS Regression: **p-value: 9e-05** R: -0.761 $R^2$: 0.58 | | | |

| | | |
|---|---|---|
| **3I:** Regression of % change of max beta power by rise in peak beta over pre-beta. | All-on Regression: p-value: 0.23 R: -0.281 $R^2$: 0.079 | RVS Regression: **p-value: 0.0003** R: -0.72 $R^2$: 0.52 |
| **Supp. 2A:** Regression of change in % change of mean beta power by rise in peak beta over pre-beta, Contralateral side only. | All-on Regression: p-value: 0.184 R: -0.412 $R^2$: 0.17 | RVS Regression: **p-value: 0.0002** R: -0.872 $R^2$: 0.76 |
| **Supp. 2B:** Regression of change in peak beta power by rise in peak beta over pre-beta, Contralateral side only. | All-on Regression: p-value: 0.76 R: -0.1 $R^2$: 0.01 | RVS Regression: **p-value: 0.006** R: -0.737 $R^2$: 0.543 |
| **Supp. 2C:** Regression of change in % change of mean beta power by rise in peak beta over pre-beta, Ipsilateral side only. | All-on Regression: p-value: 0.347 R: -0.385 $R^2$: 0.148 | RVS Regression: p-value: 0.057 R: -0.69 $R^2$: 0.48 |
| **Supp. 2D:** Regression of change in peak beta power by rise in peak beta over pre-beta, Ipsilateral side only. | All-on Regression: p-value: 0.135 R: -0.577 $R^2$: 0.332 | RVS Regression: **p-value: 0.027** R: -0.764 $R^2$: 0.583 |

Results listed for Fig 3. A, C, E, F are independent t-tests. Results for H, I, and Supp. Fig. 2 are linear regression with Wald tests for p-values. Statistical tests for D and G were LMEM derived; p-values for All-on vs. RVS. SEM is the standard error of the mean, CI is the [0.05, 0.95] confidence interval.
